# Clonal hematopoiesis is associated with protection from Alzheimer’s disease

**DOI:** 10.1101/2021.12.10.21267552

**Authors:** Hind Bouzid, Julia A. Belk, Max Jan, Yanyan Qi, Chloé Sarnowski, Sara Wirth, Lisa Ma, Matthew Chrostek, Herra Ahmad, Daniel Nachun, Winnie Yao, NHLBI Trans-omics for Precision Medicine (TOPMed) Consortium, Alexa Beiser, Alexander G. Bick, Joshua C. Bis, Myriam Fornage, William T. Longstreth, Oscar L. Lopez, Pradeep Natarajan, Bruce M. Psaty, Claudia L. Satizabal, Joshua Weinstock, Eric B. Larson, Paul K. Crane, C. Dirk Keene, Sudha Seshadri, Ansuman T. Satpathy, Thomas J. Montine, Siddhartha Jaiswal

## Abstract

Clonal hematopoiesis of indeterminate potential (CHIP) is a pre-malignant expansion of mutated blood stem cells that also associates with non-hematological disorders. Here, we tested whether CHIP was associated with Alzheimer’s disease (AD). Surprisingly, we found that CHIP carriers had reduced risk of AD dementia or AD neuropathologic features in multiple cohorts. The same mutations found in blood were also detected in the microglia-enriched fraction of brain in 7 out of 8 CHIP carriers. Single-cell chromatin accessibility profiling of brain-derived nuclei in two CHIP carriers revealed that the mutated cells were indistinguishable from microglia and comprised between 42-77% of the total microglial pool. These results suggest a role for mutant, marrow-derived cells in attenuating risk of AD, possibly by supplementing a failing microglial system during aging.

## Main Text

Hematopoietic stem cells (**HSCs**) randomly accumulate somatic mutations during aging (*1*). While most of these mutations have no consequence, rare fitness-increasing mutations may allow an HSC to clonally expand. This age-associated expansion is termed clonal hematopoiesis of indeterminate potential (**CHIP**), is found in 10-30% of those older than 70, most commonly occurs due to loss-of-function mutations in transcriptional regulators such as *DNMT3A, TET2*, and *ASXL1*, and can be detected by sequencing of DNA from peripheral blood or bone marrow cells (*2*). As these mutations are also founding mutations for hematological neoplasms such as acute myeloid leukemia, it is unsurprising that CHIP associates with increased risk of developing these cancers (*3–5*). However, CHIP also associates with increased risk of atherosclerotic cardiovascular disease and death (*6–8*). This link is believed to be causal, as mice that are deficient for *Tet2* or *Dnmt3a* in hematopoietic cells develop larger atherosclerotic plaques, presumably due to altered gene expression in mutant macrophages which favors more rapid progression of the lesions (*6, 9*).

Alzheimer’s disease (**AD**) remains a leading cause of morbidity and mortality in the aged, but therapies that can effectively slow or halt its progression are lacking. Genome-wide association studies have implicated functional alterations of microglia, the macrophage-like hematopoietic cells in the brain, as a major driver of AD risk (*10*). Given that the common CHIP-associated mutations are known to alter the development and function of myeloid cells (*6, 9, 11*), we hypothesized that CHIP may also be associated with risk of AD.

To test this hypothesis, we utilized data from the Framingham Heart Study (**FHS**) and the Cardiovascular Health Study (**CHS**), which are two cohorts within the Trans-omics for Precision Medicine (**TOPMed**) project (*12*). CHIP variants (**Table S1**) were identified from blood-derived whole genome sequencing data as previously described (*7*). Participants in CHS were substantially older on average and a higher proportion were female compared to participants in FHS (**Table S2**). The FHS subset in TOPMed included some related participants selected for family studies but was otherwise a random selection of the total cohort. The CHS subset was heavily oversampled for coronary heart disease and stroke, conditions indicative of systemic atherosclerosis (see **Materials and Methods**). Vascular dementia, which can mimic AD clinically, is thought to result from reduced blood flow in the brain in part due to atherosclerosis. Given the established association of CHIP with atherosclerotic cardiovascular disease, we excluded participants in both cohorts who had coronary heart disease or stroke. A diagnosis for AD dementia was made based on criteria from the National Institute of Neurological and Communicative Diseases and Stroke/Alzheimer’s Disease and Related Disorders Association (NINCDS/ADRDA) for definite, probable, or possible Alzheimer’s disease. We excluded anyone with a baseline diagnosis of AD dementia or missing data on incident AD dementia. After these exclusions, there were 2,437 participants in FHS, of whom 92 (3.8%) developed incident AD dementia, and 743 participants in CHS, of whom 166 (22.3%) developed incident AD dementia (**Table S2**). Since CHIP is associated with increased risk of mortality, confounding due to survivorship bias may occur when performing association studies for CHIP. To mitigate this possibility, we tested for an association between CHIP and AD dementia using competing risks regression, with death as the competing risk, while also adjusting for age, sex, *APOE* genotype, study site (for CHS), and self-reported ancestry (for CHS). Contrary to our expectations, the presence of CHIP was associated with a lower subdistribution hazard ratio (**SHR**) for incident AD dementia in both cohorts (SHR 0.69, p=0.13 in CHS; SHR 0.51, p=0.060 in FHS; SHR 0.62, p=0.021 in a fixed-effects meta-analysis of the two cohorts) (**Fig. 1A, Fig. S1**). We obtained similar results using Cox proportional hazards regression models or when including family as a clustered variable for FHS (**Table S4**).

**Fig. 1.**
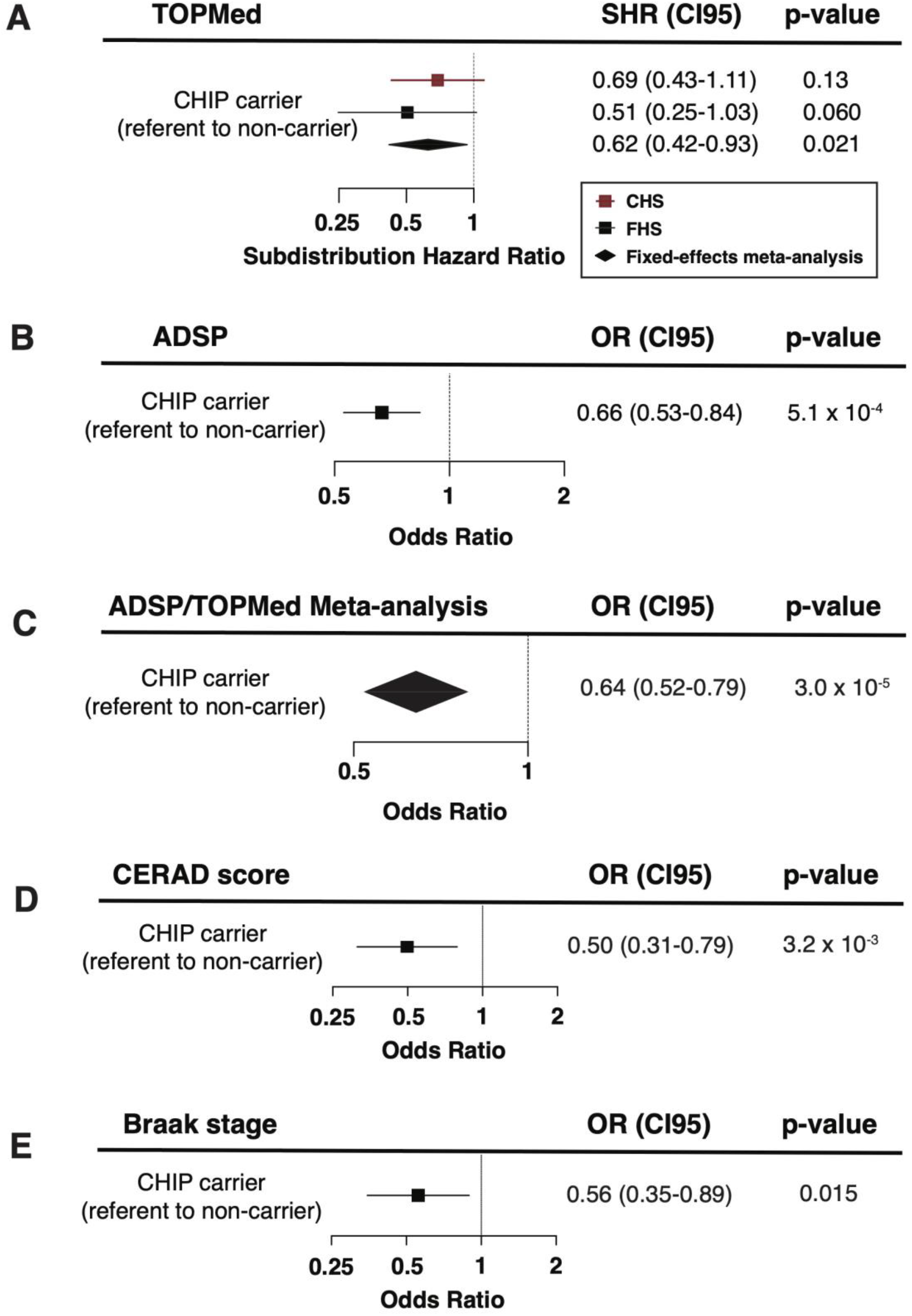
CHIP is associated with protection from AD dementia and ADNC in those without dementia. A) Forest plot for risk of incident AD in CHIP carriers from Cardiovascular Health Study (CHS) and Framingham Heart Study (FHS) relative to non-carriers. Subdistribution hazard ratios (SHR), 95% confidence intervals (CI95) and Wald p-values were calculated for each covariate from competing risks regression models which included age at blood draw, sex, and *APOE* genotype as covariates. Results from CHS and FHS were then meta-analyzed using a fixed-effects model for the two cohorts (see Figure S1 for full regression results). B) Forest plot for risk of AD in CHIP carriers relative to non-carriers from the AD Sequencing Project (ADSP) with *APOE* ε3ε3 genotype. Odds ratio (OR), 95% confidence interval (CI95) and Wald p-value were calculated from a logistic regression model that also included age at time of blood draw and sex as covariates (see Table S6 for full regression results). C) Fixed-effects meta-analysis for risk of AD in CHIP carriers using logistic regression in ADSP, FHS, and CHS. D) Plot of odds ratio (OR) and 95% confidence interval (CI95) for increased CERAD neuritic plaque score in ADSP participants without a dementia diagnosis from an ordinal logistic regression model. The covariates included in the model are age at autopsy, *APOE* genotype, sex, and CHIP status. The p-values were calculated by comparing the t-statistic for each covariate against a standard normal distribution. Full regression results are in Table S8. E) Plot of odds ratio (OR) and 95% confidence interval (CI95) for increased Braak stage in ADSP participants without a dementia diagnosis from an ordinal logistic regression model. The covariates included in the model are age at autopsy, *APOE* genotype, sex, and CHIP status. The p-values were calculated by comparing the t-statistic for each covariate against a standard normal distribution. Full regression results are in Table S8.

We next sought confirmation of this surprising inverse association in an independent cohort, the Alzheimer’s Disease Sequencing Project (**ADSP**), a case-control study for AD with whole exome sequencing (**WES**) data from brain or blood-derived DNA (*13*). *APOE* genotype is the strongest genetic risk factor for AD (*14*), with *APOE* ε2 alleles conferring protection from disease and *APOE* ε4 alleles conferring increased risk, as compared to those with *APOE* ε3ε3 (**Fig. S1**). The sample selection strategy for ADSP resulted in cases and controls that were not well-matched for age at WES blood draw for those carrying *APOE* ε2 or ε4 alleles (see **Materials and Methods**). Given CHIP’s strong association to age, this selection bias presented a major source of confounding that precluded the analysis of carriers of these alleles. However, *APOE* ε3ε3 AD dementia cases and controls were well matched for age (**Table S5**), permitting us to test for an association to CHIP in this set. After excluding those with missing information on age at blood draw, a total of 1,446 controls and 1,104 AD dementia cases with blood-derived whole exome sequencing were available for this analysis (**Table S5**). In this set, there were no overlapping participants between ADSP and TOPMed. CHIP variants were identified in ADSP using an approach previously described (*6*) and the prevalence of CHIP was appropriate for the age of the cohort (**Table S5**). The sequencing depth in ADSP was higher than for TOPMed, which resulted in greater sensitivity to detect smaller clones (**Fig. S2**). Clone size, which is approximated by the variant allele fraction (**VAF**), has previously been shown to be an important predictor of risk for blood cancer (*4, 15*) and cardiovascular outcomes (*6, 7*). In order to directly compare outcomes in ADSP to TOPMed, we limited the definition of CHIP carriers to those with VAF>0.08 in this analysis—a cutoff that was chosen because it resulted in a VAF distribution that was nearly identical to TOPMed (**Fig. S2**). We found that CHIP with VAF>0.08 was associated with reduced risk of AD dementia in ADSP (odds ratio [**OR**] 0.66, p=5.5 × 10^−4^) (**Fig. 1B, Table S6**). In contrast, having VAF≤0.08 had no association to AD dementia (OR 1.25, p=0.23) (**Table S7**), suggestive of a dose-response relationship between the size of the mutant clone and protection from AD dementia. Indeed, higher VAF was also significantly associated with protection from AD dementia when modeled as a continuous variable (**Table S7**). A meta-analysis of ADSP, CHS, and FHS showed that CHIP-carriers had a significant reduction in risk of AD (OR 0.64, p=3.0 × 10^−5^) (**Fig. 1C**). In sum, our human genetic association analyses demonstrate that CHIP is associated with protection from AD dementia in multiple cohorts, and that the degree of protection is proportional to the size of the mutant clone.

Mendelian randomization is a form of causal inference in which genetic variants known to influence the risk of a particular exposure or trait (in this case CHIP) are assessed for an association to a clinical outcome (in this case AD). We selected three independent common variants that reached genome-wide significance in a CHIP genetic association study as the instrumental variables for CHIP exposure (rs2853677, rs7726159, rs58322641)[(*7*). Summary statistics from a large AD genome-wide association study (GWAS) and GWAS-by-proxy meta-analysis(*16*) were then used to perform Mendelian randomization using the inverse-variance weighted method(*17*). Here, an increase in the genetic risk of CHIP was associated with reduced odds of AD (OR 0.92 per 1 log-odds increase in risk of CHIP, p=5.6 × 10^−3^, **Fig. S3**).

The hallmark neuropathological features of AD, regional accumulation of beta-amyloid plaques and tau neurofibrillary tangles, can also be found in some people without a clinical diagnosis of dementia. A neuritic plaque density score developed by the Consortium to Establish a Registry for AD (**CERAD**) (*18*) and Braak stage for neurofibrillary tangle distribution (*19*) are commonly used to assess for these changes at brain autopsy, with an increasing score indicative of more extensive accumulation of pathologic features. A subset of participants in ADSP had brain autopsy performed after death, which allowed us to test whether CHIP was associated with AD-related neuropathologic change (**ADNC**) in those without dementia (**Table S8**). Here, the presence of CHIP was associated with having a lower CERAD neuritic plaque score (OR 0.50, p=3.2 × 10^−3^) and Braak stage (OR 0.56, p=0.015) using ordinal logistic regression after adjusting for age, sex and *APOE* genotype (**Fig. 1D-E, Table S8**). This suggests that CHIP associates with protection from the burden of neuritic plaque and neurofibrillary tangle formation even in the absence of clinical dementia.

We next asked whether the protection from AD seen in CHIP carriers was influenced by *APOE* genotype. In those age 70 or older, *APOE* genotype was strongly associated with AD dementia risk in those without CHIP (p=5.8 × 10^−5^ by log-rank test). This effect was not seen in CHIP carriers of the same age (p=0.70 by log-rank test), though the smaller sample size in this group may have limited our power to find an association (**Fig. 2A**). In competing risks regression models stratified by *APOE* genotype, there was a similar magnitude of AD dementia risk reduction in CHIP carriers who were *APOE* ε3ε3 or who carried an *APOE* ε4 allele, but not in those who had the protective *APOE* ε2ε2 or *APOE* ε2ε3 genotypes (**Fig. 2B, Table S9**). These findings suggest that the mechanism by which CHIP associates with reduced risk of AD dementia might be redundant with the protection conferred through *APOE* ε2.

**Fig. 2.**
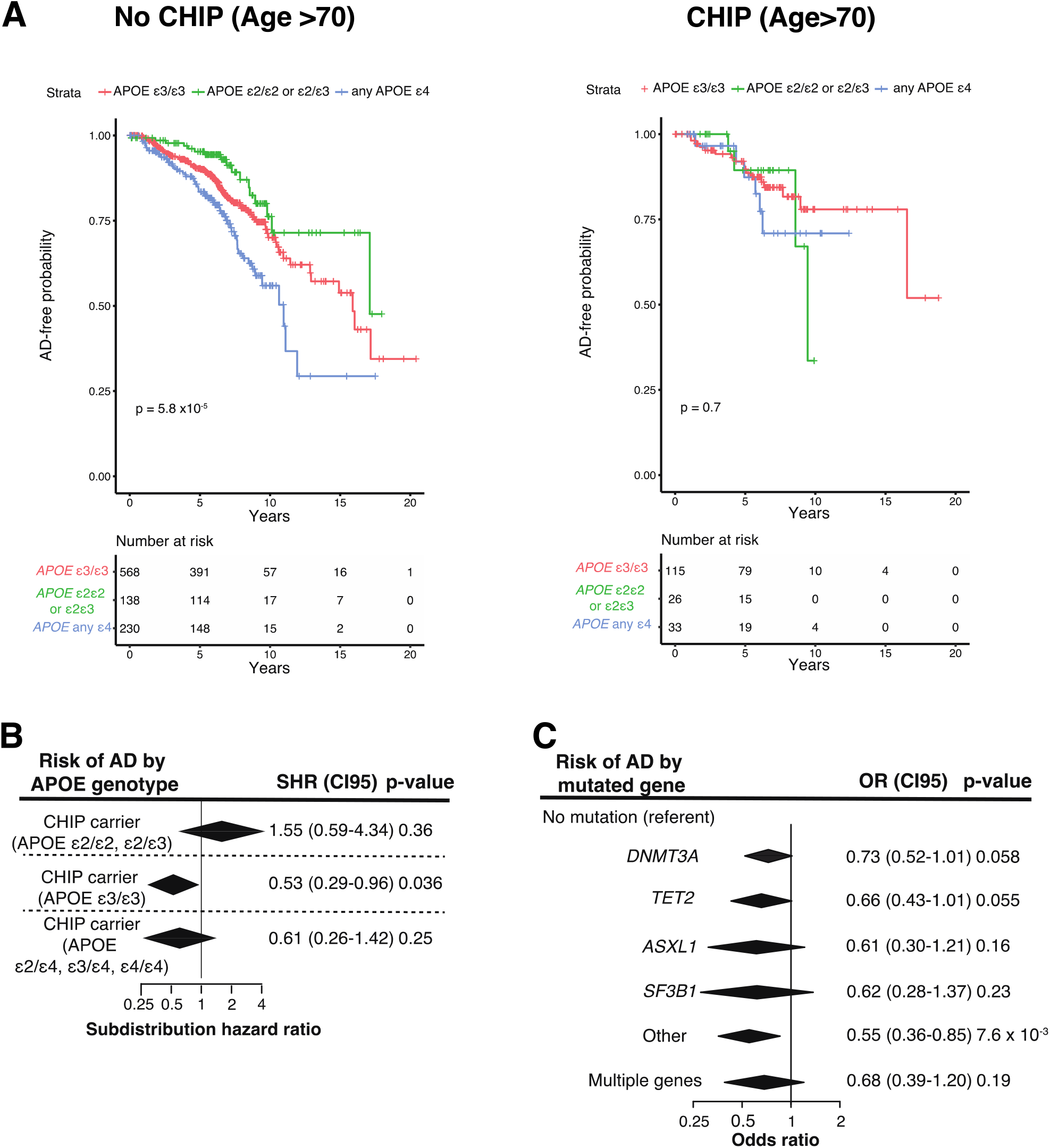
Associations of CHIP to AD by *APOE* genotype and mutated driver gene. A) Kaplan-Meier curve showing AD-free probability in CHIP non-carriers (left) and carriers (right), stratified by *APOE* genotype. Analysis was restricted to those older than 70 at time of blood draw. B) Forest plot for effect of CHIP on AD risk in participants from CHS and FHS stratified by *APOE* genotype. Participants were binned into those with neutral (*APOE* ε3ε3), low risk (*APOE* ε2ε2 and ε2ε3), and high-risk (any *APOE* ε4 allele) groups. Subdistribution hazard ratios (SHR), 95% confidence intervals (CI95) and Wald p-values were calculated for each covariate (age at time blood draw for sequencing, sex, CHIP carrier status) from competing risks regression models, and results from FHS and CHS were then meta-analyzed using a fixed-effects model (see Table S9 for full regression results). C) Plot of odds ratios for effect of mutated CHIP gene on AD in participants from the TOPMed cohorts (CHS and FHS) and ADSP. Odds ratios (OR), 95% confidence intervals (CI95) and Wald p-values were calculated for each covariate (age at time blood draw for sequencing, sex, cohort, and *APOE* genotype) from logistic regression models, and results from the TOPMed cohorts and ADSP were then meta-analyzed using a fixed-effects model (see Table S10 for full regression results).

We also assessed whether the risk of AD dementia varied based on the specific mutated gene. Of the most commonly mutated genes in CHIP, all were associated with protection from AD dementia to a similar degree (**Fig. 2C, Table S10**).

We wondered whether cells bearing CHIP-associated mutations could be found in brain, a finding that would strengthen the likelihood of a causal association between CHIP and AD risk. We obtained brain DNA-derived WES data from 1,776 persons in ADSP, of whom 1,462 had AD dementia (82.3%), and assessed for the presence of CHIP-associated variants in these samples. Similar to a prior study (*20*), we found mutations consistent with CHIP in 17 brain samples, including 15 with AD dementia (**Fig. 3A, Table S11**). Paired blood DNA was not available for the brain exome samples, so we could not determine whether the mutations we identified were indicative of blood-derived cells in brain.

**Fig. 3.**
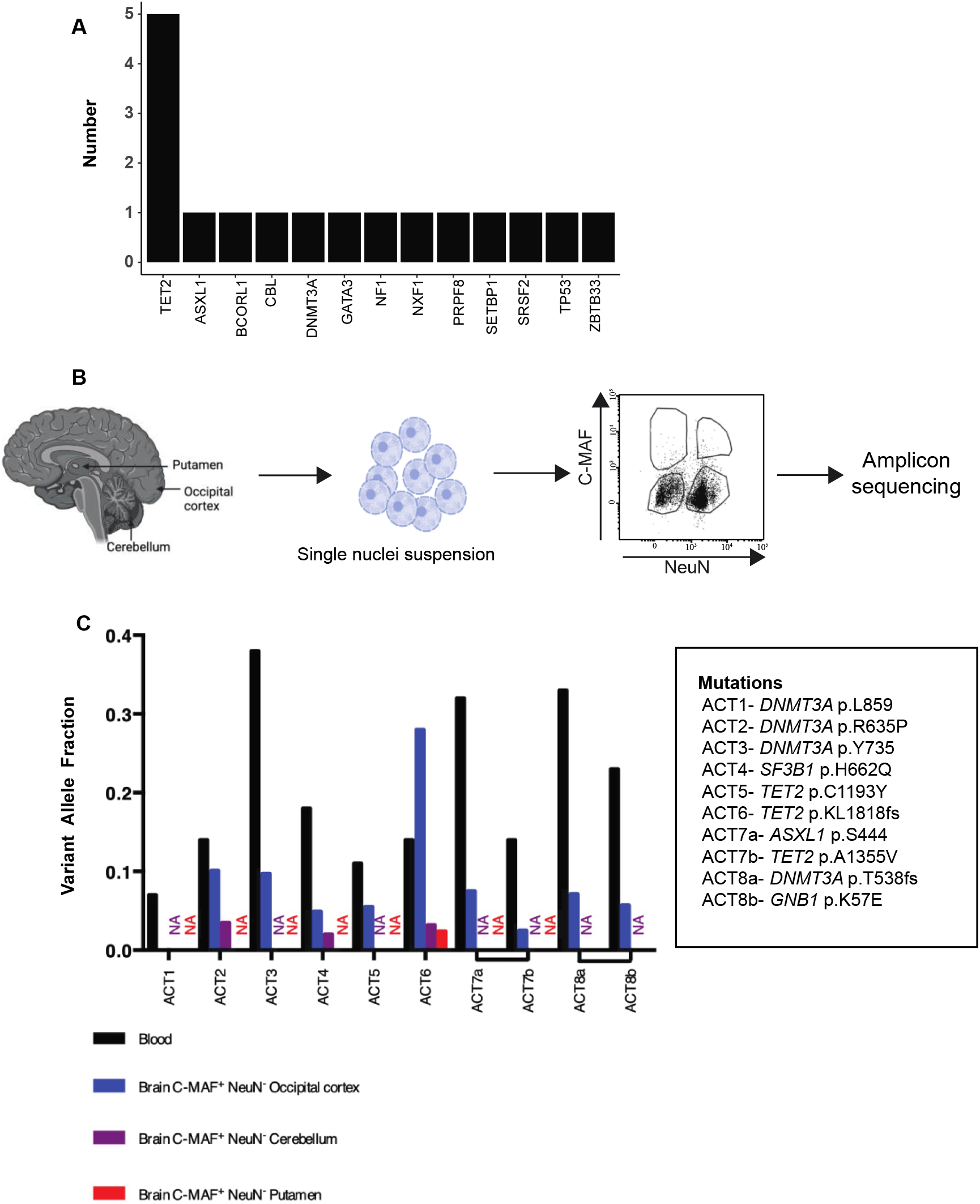
CHIP variants can be found in the microglia-enriched fraction of brain. A) Barplot of putative CHIP mutations identified from whole exome sequencing of brain DNA from 1,775 persons in ADSP. Full details on the variants identified are in Table S11. B) Schematic of experimental workflow. Autopsy samples from occipital cortex, cerebellum and putamen were digested to prepare single nuclei suspensions. Nuclei were then stained and sorted using antibodies to C-Maf^+^ (marker of myeloid cells) and NeuN+ (Marker of Neuronal cells), followed by amplicon sequencing for CHIP variants. C) Barplot of the variant allele fraction (VAF) of the CHIP variants from 8 donors (ACT1 to ACT8). For each sample, the VAF in the blood and in the brain C-Maf^+^ NeuN^-^ population are shown. Occipital cortex was available for all 8 donors. A bar for cerebellum or putamen is shown if available, otherwise NA in the corresponding color designates lack of an available sample (purple for cerebellum and red for putamen). The CHIP mutations carried by each participant are reported in the box on the right of the barplot.

Hematopoietic cells, nearly all of which are microglia, comprise ∼1-10% of the total cells in the brain across brain regions (*21*) and the limit of detection for clonal hematopoiesis by WES at 80X sequencing depth is ∼4% of cells harboring a mutation in a sample (*4*). Therefore, it would be difficult to detect clonal hematopoiesis mutations from unfractionated brain for the vast majority of CHIP carriers using WES. We hypothesized that most CHIP carriers would have the mutations detectable in the brain if examined using more sensitive methods. To test this hypothesis, we obtained tissue samples from the occipital lobe, and in some cases putamen or cerebellum, of 8 donors from the Adult Changes in Thought (**ACT**) cohort who were known to have CHIP from blood exome sequencing, as well as 1 person without CHIP (**Table S12**). All persons were in their 80s and 7 out of 9 were without dementia and had no/low ADNC at the time of death. The 8 CHIP carriers had mutations in *DNMT3A, TET2, ASXL1, SF3B1*, and *GNB1* with the highest frequency in *DNMT3A* (4 out of 8) and *TET2* (3 out of 8) (**Table S12**), which is representative of the relative proportion of these mutations in the general population (*7*). In addition, 2 out of the 8 harbored two different CHIP mutations. To determine whether bone marrow derived cells carrying CHIP mutations were present in the brains of these individuals, we digested the frozen brain tissue and isolated intact nuclei, from which we extracted DNA for amplicon sequencing. We detected the same mutations that were present in blood in 6 out of 8 unfractionated brains with VAF ranging from 0.004 to 0.02 (**Fig. S4, Table S12**). Of note, the two donors where CHIP variants were not identified in unfractionated brain (ACT1, ACT8) both had AD dementia during life. In contrast, all 6 donors where CHIP variants were found in unfractionated brain were without dementia and free of ADNC.

The CHIP variants detected in whole brain DNA might have originated from residual circulating hematopoietic cells in the vasculature, such as granulocytes or lymphocytes. Alternatively, myeloid cells such as microglia in the brain parenchyma could have been the source of the mutations, although microglia are believed to have little contribution from HSC-derived cells in adulthood (*22*). To distinguish between these possibilities, we conceived a strategy to enrich for these cells from frozen brain tissue. Since the tissue was not viably cryopreserved, isolation of cells based on expression of membrane antigens was not possible. Instead, we used antibodies to nuclear transcription factors to enrich for mononuclear phagocytes, such as macrophages and microglia **(Fig. 3B)**. We stained nuclei for the neuronal-specific transcription factor NeuN (RBFOX3) and c-Maf, a transcription factor expressed in phagocytes as well as some neurons and non-hematopoietic glial cells. We then sorted 4 populations based on the presence or absence of these markers **(Fig. 3B)**. The CHIP somatic variants were found in the NeuN^-^ c-Maf^+^ population in 7 out 8 brains, with a VAF that ranged from 0.02 to 0.28 (representing 4% to 56% of NeuN^-^ c-Maf^+^ nuclei). In contrast, CHIP somatic variants were not detected in the NeuN+ c-Maf-neuronal population and were absent or at low levels in the other two populations (**Fig. 3C, Fig. S4)**. The VAF was lower in the NeuN-c-Maf+ nuclei from cerebellum compared to occipital cortex in the three samples where tissue from both regions was available. In sample ACT8, CHIP variants were robustly detected in the NeuN-c-Maf+ population from occipital cortex but not putamen. These results indicate that there is a substantial contribution to the brain mononuclear phagocytic pool from circulating mutated cells, and that there is also regional heterogeneity in the frequency of infiltrating CHIP+ cells in brain.

Our flow cytometric analysis indicated that a prominent myeloid population bearing CHIP mutations was present in the brains of most CHIP carriers. However, it was unknown if the mutated cells were similar to endogenous brain microglia or were instead a distinct myeloid population not normally found in brain, such as monocyte-derived macrophages or dendritic cells. To better understand the phenotype of these mutated cells, we performed single-cell assay for transposase-accessible chromatin sequencing (**scATAC-seq**) on brain samples from three ACT participants: a brain donor without CHIP (ACT9), one with *TET2*-mutant CHIP (ACT6), and one with *DNMT3A*-mutant CHIP (ACT2) (**Table S12**). For the ACT6 donor, we analyzed tissue from cerebellum and putamen, whereas occipital cortex was assessed for the other two donors. scATAC-seq was performed on unsorted nuclei, as well as sorted NeuN-c-Maf+ nuclei for each sample. After aligning and filtering the scATAC-seq reads, our samples had a median of 12,287 fragments per cell and a median enrichment of fragments in transcription start sites of 9.31, indicating that we recovered high quality scATAC-seq libraries from these archived samples (**Fig. S5**). In total, we recovered high quality scATAC-seq profiles for 38,206 cells. We then aggregated our data with scATAC-seq data from 10 samples (an additional 72,984 cells) from a comprehensive scATAC-seq characterization of the adult human brain (Corces 2020) (*21*). After clustering and dimensionality reduction, we identified 18 clusters encompassing the major brain cell types (**Fig. 4A-B**), including one cluster that contained previously described microglia as well as myeloid cells from each of our samples (cluster 9) (**Fig. 4B, Fig. S6, Table S13**). No other hematopoietic cell types were observed in the human brain samples. For cells within cluster 9 grouped by sample, inspecting pseudo-bulk ATAC-seq tracks revealed accessible chromatin at the microglia marker genes *TMEM119, P2RY12, SALL1, CSF1R*, and *SIGLEC8* in each of our samples, which was visually similar to the reference microglia. Cells in this cluster also had high accessibility at the AD-associated genes *APOE, TREM2, TYROBP, AXL*, and *MERTK* (**Fig. 4C, Fig. S6**). As an additional control, we examined reference scATAC-seq profiles of blood monocytes and classical dendritic cells (cDCs) (*23*) and found that these cells had little to no accessibility at the aforementioned microglia genes. Furthermore, the Cluster 9 tracks in all brain samples had low accessibility at *ANPEP* (CD13), *CXCL2*, and *ITGAE* (CD103), in contrast to monocytes or cDCs (**Fig. 4C, Fig. S6**). Finally, we quantified the genome-wide similarity of microglia in each pair of samples by considering the number of differential peaks between each pair for cells within cluster 9. The putamen-derived sample showed modest differences from the occipital cortex-derived samples but otherwise essentially no differences were observed, and all comparisons were within the range of variation observed when comparing pairs of the Corces 2020 samples (**Fig. S6**). These results indicate that the cells in cluster 9 are indistinguishable from microglia and unlikely to contain contaminating monocytes or dendritic cells.

**Fig. 4.**
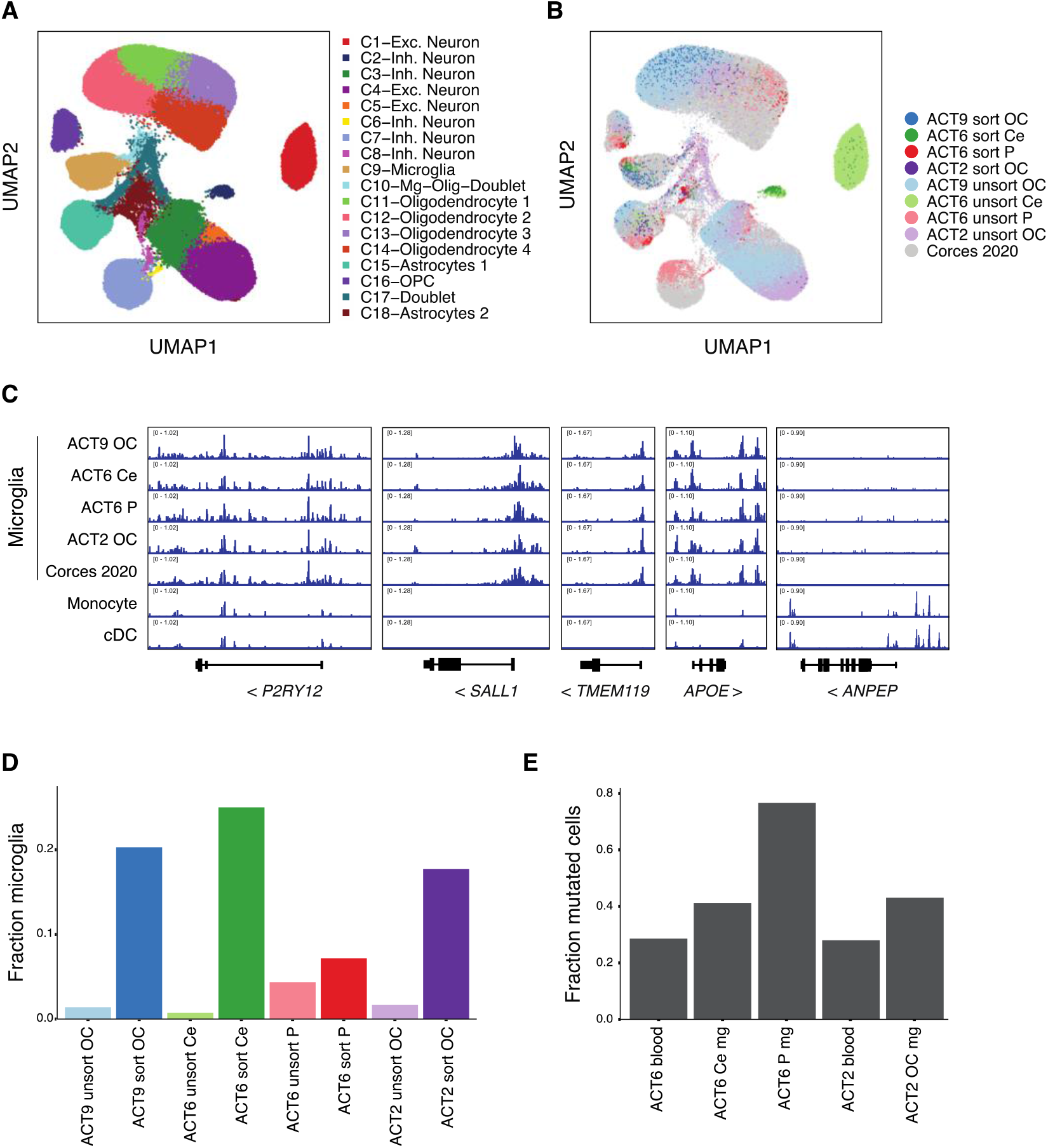
scATAC-seq of brain samples from CHIP carriers reveals that the mutated cells are similar to microglia and comprise a large proportion of the microglial pool. A) scATAC-seq profiles of 111,190 cells from our dataset and the Corces 2020 adult human brain dataset. Each dot represents the scATAC-seq profile of one cell and is colored by its assigned cluster. B) scATAC-seq profiles of all cells colored by which sample it originated from. Samples from Corces 2020 are aggregated and shown in grey. Sorted samples were from the c-Maf+ NeuN-gate. C) Pseudo-bulk tracks for selected gene loci. The top 5 tracks show scATAC-seq coverage of cells from the indicated sample (or aggregated Corces 2020 samples) within C9, the microglia cluster. The monocyte and classical dendritic cell (cDC) tracks are from the Satpathy 2019 hematopoiesis dataset. D) Fraction of cells in cluster C9 (microglia) for sorted versus unsorted brain samples. E) Proportion of microglia (mg) bearing a CHIP mutation in each sample, calculated by dividing the percentage of cells in cluster 9 in each unsorted sample by 2 times the VAF of the CHIP mutation for that sample. The brain regions are abbreviated as Ce for Cerebellum, OC for Occipital cortex, and P for putamen.

Having established that the only hematopoietic cell type present in these brains was microglia, we used the scATAC-seq data to evaluate the effectiveness of our flow cytometric method for enrichment of these cells. We compared the proportion of microglia in the sorted samples to those present in the unsorted samples and observed that microglia were enriched 11 to 33-fold in the sorted samples from occipital cortex and cerebellum, but only modestly in the putamen sample (**Fig. 4D**). The percentage of microglia ranged from 7.2% to 25% in the sorted samples, indicating that there were still large numbers of contaminating non-microglial cells in the NeuN-c-Maf+ gate. This suggested that the true fraction of mutated microglia was substantially underestimated by our previous approach using amplicon sequencing of NeuN-c-Maf+ nuclei. To estimate the percentage of mutated microglia more accurately, we first assessed the VAF for the CHIP variant in each unsorted brain sample. Since these are heterozygous mutations, multiplying the VAF by 2 gives an estimate of the percentage of mutated cells in the sample. Since microglia were the only hematopoietic cell type present in these brains, we reasoned that we could divide the percentage of total mutated cells by the percentage of microglia in each unsorted sample to estimate the percentage of mutant microglia. Using this approach, we calculated that 43% of the microglia in ACT2 harbored the *DNMT3A* mutation, as compared to 28% of circulating cells in the blood. For the ACT6 donor, 77% and 42% of putamen and cerebellar microglia harbored the *TET2* mutation, respectively, compared to 28% of circulating blood cells (**Fig. 4E, Table S14**). These results indicate that replacement of endogenous microglia by mutant, marrow-derived cells is widespread in the aging brain. The observation that the proportion of mutated cells is substantially greater in the microglial pool than in the blood also suggests that there is positive selection for the mutant cells in the brain microenvironment.

We show here that, unexpectedly, the presence of CHIP is associated with protection from AD dementia. This effect is seen in multiple cohorts, is not due to survival bias, is seen with several different mutated genes, and is strongest in carriers of *APOE* ε3ε3 or *APOE* ε4 alleles. The degree of protection from AD dementia seen in CHIP carriers is greater than carrying an *APOE* ε2 allele, which is the most protective common inherited variant for AD (*24*). CHIP is also associated with lower levels of neuritic plaques and neurofibrillary tangles in those without dementia, indicating a possible modulating effect of CHIP on the underlying pathophysiology of AD. Consistent with this hypothesis, we also detect substantial infiltration of brain by marrow-derived mutant cells which adopt a microglial-like phenotype. We speculate that the mutations associated with CHIP confer circulating precursor cells with an enhanced ability to engraft in the brain, to differentiate into microglia once engrafted, and/or to clonally expand relative to unmutated cells in the brain microenvironment. These non-mutually exclusive possibilities could provide protection from AD by supplementing the phagocytic capacity of the endogenous microglial system during aging. Alternatively, or in addition, the mutations may alter the functionality of the engrafted myeloid cells in a manner that promotes clearance of pathologic beta-amyloid and/or tau. Understanding the interplay between CHIP and the aging brain may yield valuable information about the pathogenesis of AD and provide insights into slowing its progression.

## Data Availability

All data from TOPMed and ADSP are available on dbGaP for investigators with approved protocols.
Single-cell ATAC-seq data generated here from human brain samples will be deposited in Gene Expression Omnibus.

## Acknowledgements

We thank Marilynn Miller, Kelley Faber, Amanda Kuzma, William Lee and KatieRose Richmire for providing age at blood draw for ADSP participants, Aimee Schantz for compiling the neuropathology data from ACT, Walter Kukull and Jessica Culhane for compiling neuropathology data from NACC/ADRCs, and Ben Ebert and Ryan Corces for helpful discussions. Most importantly, we thank the studies and participants who provided biological samples and data.

The following are study specific acknowledgements:

## TOPMed

WGS for the TOPMed program was supported by the National Heart, Lung and Blood Institute (NHLBI). Centralized read mapping and genotype calling, along with variant quality metrics and filtering were provided by the TOPMed Informatics Research Center (3R01HL-117626-02S1; contract HHSN268201800002I). Phenotype harmonization, data management, sample-identity quality control and general study coordination were provided by the TOPMed Data Coordinating Center (R01HL-120393; U01HL-120393; contract HHSN268201800001I). Genome sequencing for “NHLBI TOPMed: Whole Genome Sequencing and Related Phenotypes in the Framingham Heart Study” (phs000974.v1.p1) was performed at the Broad Institute Genomics Platform (3R01HL092577-06S1, 3U54HG003067-12S2). Genome sequencing for “NHLBI TOPMed: Whole Genome Sequencing Project: Cardiovascular Health Study” (phs001369.v2.p2) was performed at the Broad Institute Genomics Platform (3R01HL092577-06S1, 3U54HG003067-12S2) and Baylor University (HHSM268201600033I, 3U54HG003273-12S2, HHSN268201500015C). We gratefully acknowledge the studies and participants who provided biological samples and data for TOPMed.

Framingham Heart Study: The Framingham Heart Study (FHS) acknowledges the support of contracts NO1-HC-25195, HHSN268201500001I and 75N92019D00031 from the National Heart, Lung and Blood Institute and grant supplement R01 HL092577-06S1 for this research. We also acknowledge the dedication of the FHS study participants without whom this research would not be possible. Dr. Vasan is supported in part by the Evans Medical Foundation and the Jay and Louis Coffman Endowment from the Department of Medicine, Boston University School of Medicine.

The FHS was also supported by grants from the National Institutes of Aging (AG054076 and AG058589).

Cardiovascular Health Study: This research was supported by contracts HHSN268201200036C, HHSN268200800007C, HHSN268201800001C, N01HC55222, N01HC85079, N01HC85080, N01HC85081, N01HC85082, N01HC85083, N01HC85086, 75N92021D00006, and grants U01HL080295 and U01HL130114 from the National Heart, Lung, and Blood Institute (NHLBI), with additional contribution from the National Institute of Neurological Disorders and Stroke (NINDS). Additional support was provided by R01AG023629 from the National Institute on Aging (NIA). A full list of principal CHS investigators and institutions can be found at CHS-NHLBI.org. The content is solely the responsibility of the authors and does not necessarily represent the official views of the National Institutes of Health.

## ADSP

The Alzheimer’s Disease Sequencing Project (ADSP) is comprised of two Alzheimer’s Disease (AD) genetics consortia and three National Human Genome Research Institute (NHGRI) funded Large Scale Sequencing and Analysis Centers (LSAC). The two AD genetics consortia are the Alzheimer’s Disease Genetics Consortium (ADGC) funded by NIA (U01 AG032984), and the Cohorts for Heart and Aging Research in Genomic Epidemiology (CHARGE) funded by NIA (R01 AG033193), the National Heart, Lung, and Blood Institute (NHLBI), other National Institute of Health (NIH) institutes and other foreign governmental and non-governmental organizations. The Discovery Phase analysis of sequence data is supported through UF1AG047133 (to Drs. Schellenberg, Farrer, Pericak-Vance, Mayeux, and Haines); U01AG049505 to Dr. Seshadri; U01AG049506 to Dr. Boerwinkle; U01AG049507 to Dr. Wijsman; and U01AG049508 to Dr. Goate and the Discovery Extension Phase analysis is supported through U01AG052411 to Dr. Goate, U01AG052410 to Dr. Pericak-Vance and U01 AG052409 to Drs. Seshadri and Fornage. Data generation and harmonization in the Follow-up Phases is supported by U54AG052427 (to Drs. Schellenberg and Wang).

The ADGC cohorts include: Adult Changes in Thought (ACT) funded by NIA (U01AG006781 and U19AG066567), the Alzheimer’s Disease Research Centers (ADRCs) including the University of Washington ADRC funded by NIA (P50AG005136 and P30AG066509), the Chicago Health and Aging Project (CHAP), the Memory and Aging Project (MAP), Mayo Clinic (MAYO), Mayo Parkinson’s Disease controls, University of Miami, the Multi-Institutional Research in Alzheimer’s Genetic Epidemiology Study (MIRAGE), the National Cell Repository for Alzheimer’s Disease (NCRAD), the National Institute on Aging Late Onset Alzheimer’s Disease Family Study (NIA-LOAD), the Religious Orders Study (ROS), the Texas Alzheimer’s Research and Care Consortium (TARC), Vanderbilt University/Case Western Reserve University (VAN/CWRU), the Washington Heights-Inwood Columbia Aging Project (WHICAP) and the Washington University Sequencing Project (WUSP), the Columbia University Hispanic-Estudio Familiar de Influencia Genetica de Alzheimer (EFIGA), the University of Toronto (UT), and Genetic Differences (GD).

The CHARGE cohorts are supported in part by National Heart, Lung, and Blood Institute (NHLBI) infrastructure grant HL105756 (Psaty), RC2HL102419 (Boerwinkle) and the neurology working group is supported by the National Institute on Aging (NIA) R01 grant AG033193. The CHARGE cohorts participating in the ADSP include the following: Austrian Stroke Prevention Study (ASPS), ASPS-Family study, and the Prospective Dementia Registry-Austria (ASPS/PRODEM-Aus), the Atherosclerosis Risk in Communities (ARIC) Study, the Cardiovascular Health Study (CHS), the Erasmus Rucphen Family Study (ERF), the Framingham Heart Study (FHS), and the Rotterdam Study (RS). ASPS is funded by the Austrian Science Fond (FWF) grant number P20545-P05 and P13180 and the Medical University of Graz. The ASPS-Fam is funded by the Austrian Science Fund (FWF) project I904), the EU Joint Programme - Neurodegenerative Disease Research (JPND) in frame of the BRIDGET project (Austria, Ministry of Science) and the Medical University of Graz and the Steiermärkische Krankenanstalten Gesellschaft. PRODEM-Austria is supported by the Austrian Research Promotion agency (FFG) (Project No. 827462) and by the Austrian National Bank (Anniversary Fund, project 15435. ARIC research is carried out as a collaborative study supported by NHLBI contracts (HHSN268201100005C, HHSN268201100006C, HHSN268201100007C, HHSN268201100008C, HHSN268201100009C, HHSN268201100010C, HHSN268201100011C, and HHSN268201100012C). Neurocognitive data in ARIC is collected by U01 2U01HL096812, 2U01HL096814, 2U01HL096899, 2U01HL096902, 2U01HL096917 from the NIH (NHLBI, NINDS, NIA and NIDCD), and with previous brain MRI examinations funded by R01-HL70825 from the NHLBI. CHS research was supported by contracts HHSN268201200036C, HHSN268200800007C, N01HC55222, N01HC85079, N01HC85080, N01HC85081, N01HC85082, N01HC85083, N01HC85086, and grants U01HL080295 and U01HL130114 from the NHLBI with additional contribution from the National Institute of Neurological Disorders and Stroke (NINDS). Additional support was provided by R01AG023629, R01AG15928, and R01AG20098 from the NIA. FHS research is supported by NHLBI contracts N01-HC-25195 and HHSN268201500001I. This study was also supported by additional grants from the NIA (R01s AG054076, AG049607 and AG033040 and NINDS (R01 NS017950). The ERF study as a part of EUROSPAN (European Special Populations Research Network) was supported by European Commission FP6 STRP grant number 018947 (LSHG-CT-2006-01947) and also received funding from the European Community’s Seventh Framework Programme (FP7/2007-2013)/grant agreement HEALTH-F4-2007-201413 by the European Commission under the programme “Quality of Life and Management of the Living Resources” of 5th Framework Programme (no. QLG2-CT-2002-01254). High-throughput analysis of the ERF data was supported by a joint grant from the Netherlands Organization for Scientific Research and the Russian Foundation for Basic Research (NWO-RFBR 047.017.043). The Rotterdam Study is funded by Erasmus Medical Center and Erasmus University, Rotterdam, the Netherlands Organization for Health Research and Development (ZonMw), the Research Institute for Diseases in the Elderly (RIDE), the Ministry of Education, Culture and Science, the Ministry for Health, Welfare and Sports, the European Commission (DG XII), and the municipality of Rotterdam. Genetic data sets are also supported by the Netherlands Organization of Scientific Research NWO Investments (175.010.2005.011, 911-03-012), the Genetic Laboratory of the Department of Internal Medicine, Erasmus MC, the Research Institute for Diseases in the Elderly (014-93-015; RIDE2), and the Netherlands Genomics Initiative (NGI)/Netherlands Organization for Scientific Research (NWO) Netherlands Consortium for Healthy Aging (NCHA), project 050-060-810. All studies are grateful to their participants, faculty and staff. The content of these manuscripts is solely the responsibility of the authors and does not necessarily represent the official views of the National Institutes of Health or the U.S. Department of Health and Human Services.

The four LSACs are: the Human Genome Sequencing Center at the Baylor College of Medicine (U54 HG003273), the Broad Institute Genome Center (U54HG003067), The American Genome Center at the Uniformed Services University of the Health Sciences (U01AG057659), and the Washington University Genome Institute (U54HG003079). Biological samples and associated phenotypic data used in primary data analyses were stored at Study Investigators institutions, and at the National Cell Repository for Alzheimer’s Disease (NCRAD, U24AG021886) at Indiana University funded by NIA. Associated Phenotypic Data used in primary and secondary data analyses were provided by Study Investigators, the NIA funded Alzheimer’s Disease Centers (ADCs), and the National Alzheimer’s Coordinating Center (NACC, U01AG016976) and the National Institute on Aging Genetics of Alzheimer’s Disease Data Storage Site (NIAGADS, U24AG041689) at the University of Pennsylvania, funded by NIA, and at the Database for Genotypes and Phenotypes (dbGaP) funded by NIH. This research was supported in part by the Intramural Research Program of the National Institutes of health, National Library of Medicine. Contributors to the Genetic Analysis Data included Study Investigators on projects that were individually funded by NIA, and other NIH institutes, and by private U.S. organizations, or foreign governmental or nongovernmental organizations.

## NACC

The NACC database is funded by NIA/NIH Grant U24 AG072122. NACC data are contributed by the NIA-funded ADCs: P30 AG019610 (PI Eric Reiman, MD), P30 AG013846 (PI Neil Kowall, MD), P50 AG008702 (PI Scott Small, MD), P50 AG025688 (PI Allan Levey, MD, PhD), P50 AG047266 (PI Todd Golde, MD, PhD), P30 AG010133 (PI Andrew Saykin, PsyD), P50 AG005146 (PI Marilyn Albert, PhD), P50 AG005134 (PI Bradley Hyman, MD, PhD), P50 AG016574 (PI Ronald Petersen, MD, PhD), P50 AG005138 (PI Mary Sano, PhD), P30 AG008051 (PI Thomas Wisniewski, MD), P30 AG013854 (PI Robert Vassar, PhD), P30 AG008017 (PI Jeffrey Kaye, MD), P30 AG010161 (PI David Bennett, MD), P50 AG047366 (PI Victor Henderson, MD, MS), P30 AG010129 (PI Charles DeCarli, MD), P50 AG016573 (PI Frank LaFerla, PhD), P50 AG005131 (PI James Brewer, MD, PhD), P50 AG023501 (PI Bruce Miller, MD), P30 AG035982 (PI Russell Swerdlow, MD), P30 AG028383 (PI Linda Van Eldik, PhD), P30 AG053760 (PI Henry Paulson, MD, PhD), P30 AG010124 (PI John Trojanowski, MD, PhD), P50 AG005133 (PI Oscar Lopez, MD), P50 AG005142 (PI Helena Chui, MD), P30 AG012300 (PI Roger Rosenberg, MD), P30 AG049638 (PI Suzanne Craft, PhD), P50 AG005136 (PI Thomas Grabowski, MD), P50 AG033514 (PI Sanjay Asthana, MD, FRCP), P50 AG005681 (PI John Morris, MD), P50 AG047270 (PI Stephen Strittmatter, MD, PhD).

## Author contributions

H.B. conceptualized experimental design and performed all research investigations on brain tissue. J.A.B. performed single-cell ATAC-seq experiments and analysis with assistance from Y.Q. and W.Y. M.J. performed analyses and data curation of ADSP data. S.W., L.M., M.C., H.A., and D.N. provided genotyping and variant calling for CHIP from human samples. A.B., J.C.B., M.F., W.T.L., O.L.L., B.M.P., C.L.S., and S.S. contributed to neurocognitive phenotype curation in CHS and FHS and provided guidance on epidemiological study design. A.G.B., P.N., and J.W. performed CHIP calling in TOPMed. E.B.L., P.K.C., and C.D.K. provided brain tissues and neuropathology data from ACT. A.T.S. provided analysis, resources, and supervision for single-cell ATAC-seq. T.J.M. provided supervision, conceptualization, and access to resources for studies involving brain autopsy tissue. S.J. conceived the research plan and supervision for all analyses. H.B., J.A.B., and S.J. wrote the original draft with review from all authors

## Data and materials availability

All genetic data from TOPMed and ADSP are available on dbGaP for investigators with approval protocols. Single-cell ATAC-seq data from human brain samples will be deposited in Gene Expression Omnibus.

