## Supplementary File for "Clonal hematopoiesis is associated with protection from Alzheimer’s disease"

**This PDF file includes:**

Materials and Methods
Figs. S1 to S6
Tables S1 to S14

#### Materials and Methods

##### Cohort descriptions

*Framingham Heart Study*

The FHS is a single-site, prospective and population-based study that has followed participants from the town of Framingham, MA to investigate risk factors for cardiovascular diseases. The population of Framingham was almost entirely white at the beginning of the study. The FHS comprises three generations of participants. The first generation (Original cohort/Gen1)(*25*), followed since 1948, enrolled 5,209 men and women who comprised two-thirds of the adult population then residing in Framingham, MA, USA. Survivors continue to receive biennial examinations. The second generation (Offspring cohort/Gen2)(*26*), followed since 1971, comprised 5,124 offspring of Gen1 and spouses of the offspring (including 3,514 biological offspring) who attended examinations every 4 to 8 years. The third generation (Gen3)(*27*) , enrolled in 2002, included 4,095 children from the largest offspring families who attended three examinations 4 years apart. All cohorts continue under active surveillance for cardiovascular events, stroke, and dementia. All participants provided written informed consent at each examination.

A total of 4,195 samples were sequenced as part of the Trans-omics for Precision Medicine (TOPMed) project Freeze 6 release as previously described (*7*). The selection of participants for sequencing was mostly a random selection of those with available DNA, but also included some related individuals for family studies. After exclusion of participants with coronary heart disease, ischemic stroke, or with missing information on age at blood draw or Alzheimer's disease (AD) diagnosis, a total of 2,437 persons remained for this analysis. Adjudication for Alzheimer's phenotypes was done by a committee (*28*), comprising at least two neurologists and a neuropsychologist. Multiple types of information were used to evaluate participants with suspected dementia, including neurologic and neuropsychological assessments, a telephone interview with a family member or care giver, medical records, imaging studies, and autopsy data when available. AD was diagnosed when participants met the criteria of the National Institute of Neurological and Communicative Disorders and Stroke (**NINCDS**) and the Alzheimer's Disease and Related Disorders Association (**ADRDA**) for definite, probable, or possible Alzheimer's disease.

Data for FHS are available in dbGaP:
https://www.ncbi.nlm.nih.gov/projects/gap/cgi-bin/study.cgi?study_id=phs000974.v1.p1

*Cardiovascular Health Study*

The CHS is a prospective, multi-ethnic, longitudinal study of risk factors for coronary heart disease and stroke in people aged 65 and older. A total of 2,840 samples were sequenced as part of the Trans-omics for Precision Medicine (TOPMed) project as previously described (*7*).The samples selected for whole genome sequencing as part of TOPMed were heavily oversampled for cardiovascular disease cases. After exclusion of participants with coronary heart disease or stroke, or with missing information on age at blood draw or AD diagnosis, a total of 743 persons remained for this analysis. Alzheimer's disease was diagnosed as probable and possible following the NINCDS-ADRDA criteria in 1997-98 and 2002-03 (*29*).

Data for CHS are available in dbGaP:
https://www.ncbi.nlm.nih.gov/projects/gap/cgi-bin/study.cgi?study_id=phs001368.v2.p2

Written informed consent was obtained from all human participants by each of the studies that contributed to TOPMed. Each study received institutional certification before deposition in dbGaP, which certified that all relevant institutional ethics committees approved the individual studies and that the genomic and phenotypic data submission was compliant with all relevant ethical regulations. Secondary analysis of the dbGaP data in this manuscript was approved by the Stanford University Institutional Review Board, and this work is compliant with all relevant ethical regulations.

*ADSP*

The Alzheimer's Disease Sequencing Project (ADSP) is a collaborative effort of the National Institutes of Aging, the National Human Genome Research Institute, and the Alzheimer's community to understand the genetic basis of AD (*13*). The whole exome sequencing (WES) set of ADSP was a case-control design where cases met NINCDS-ADRDA criteria for possible, probable, or definite AD, had documented age at onset or age at death, and *APOE* genotyping. A case-control selection strategy was chosen that targeted cases with minimal risk as predicted by known risk factors (age, sex, and *APOE*) and targeted controls with the least probability of conversion to AD by age 85 years (*13*). A total of 5,096 cases and 4,965 controls from 24 cohorts were chosen for WES. As a result of this selection strategy, cases and controls were not well-matched for age, except for carriers of *APOE* ε3ε3 genotype. Furthermore, 1,776 samples were sequenced from brain, not blood, DNA. We limited our CHIP/AD association analysis to those samples where DNA was derived from blood and where age at blood draw used for sequencing was known. In most cases, the AD diagnosis was made prior to the blood draw, however the diagnosis was usually within 5 years of the time of blood sampling for both prevalent and incident cases. After excluding those without blood DNA or known age at blood draw and further limiting to *APOE* ε3ε3 carriers, we had 1,104 AD cases and 1,446 controls who were well matched by age.

Written informed consent was obtained from all human participants by each of the studies that contributed to ADSP. Secondary analysis of the dbGaP data in this manuscript was approved by the Partners Healthcare Institutional Review Board, and this work is compliant with all relevant ethical regulations.

Data for ADSP is available in dbGaP:

https://www.ncbi.nlm.nih.gov/projects/gap/cgi-bin/study.cgi?study_id=phs000572.v8.p4

**Variant calling and annotation**

Whole genome sequencing for TOPMed samples was performed as previously described (*12*). FASTQ files were aligned to hg38 for TOPMed WGS and hg19 for ADSP and the resulting BAM files were passed through Mutect/Indelocator (ADSP) or Mutect2 (TOPMed) pipelines to identify putative variants as previously described (*6*). Briefly, the Mutect/Mutect2 pipelines excluded variants that had characteristics of common artifacts, such as oxoguanine artifact, end of read artifact, and PCR artifact (strand bias). Common polymorphisms present in germline databases were also excluded. Rare error modes were excluded by using a Panel of Normals compiled from persons without CHIP in the same sequencing centers. Output from the Mutect/Mutect2 pipelines were then annotated for known CHIP variants in 73 genes from a curated whitelist (Supplemental Table 1).

**Statistical analysis plan**

*TOPMed*

We wished to test for an association of AD dementia to CHIP. We hypothesized that CHIP carriers would have increased risk of AD dementia based on prior data that CHIP carriers have more inflammation in innate immune cells (*6*, *7*, *9*) and that enhanced inflammasome activation was associated with worsened AD phenotypes in mice (*30*).

For the discovery set, we utilized the two cohorts in TOPMed, FHS and CHS. The CHS sample was heavily oversampled for those with cardiovascular diseases, especially coronary heart disease (CHD) and stroke (1,838 out of 2,840 participants had these conditions). CHIP is known to be associated with atherosclerotic cardiovascular disease (*6*). As systemic atherosclerosis is a risk factor for vascular dementia, which can mimic AD dementia symptoms, we wished to exclude anyone with these conditions to prevent confounding. To do this, we excluded anyone with an event type of myocardial infarction (MI), stroke, angioplasty, coronary artery bypass surgery, silent MI, or death due to coronary heart disease using CHS event codes (https://www.ncbi.nlm.nih.gov/projects/gap/cgi-bin/variable.cgi?study_id=phs000287.v2.p1&phv=100824&phd=2793&pha=3548&pht=1466&phvf=&phdf=&phaf=&phtf=&dssp=1&consent=&temp=1) and excluded those selected on the basis of CHD, stroke, or "other" sampling group codes (https://www.ncbi.nlm.nih.gov/projects/gap/cgi-bin/variable.cgi?study_id=phs001368.v2.p2&phv=377683&phd=8024&pha=&pht=7957&phvf=&phdf=&phaf=&phtf=&dssp=1&consent=&temp=1). For FHS, we also excluded anyone with codes for coronary heart disease or ischemic stroke.

FHS and CHS are both prospective studies with information on incident AD diagnosis. We therefore utilized regression models to test for an association of CHIP to incident AD dementia in both cohorts. After excluding those without information on AD diagnosis, there were 2,437 persons in FHS and 620 persons in CHS. Other variables included in these models were age at blood draw used for sequencing, *APOE* genotype, and sex. The results from both cohorts were then meta-analyzed using a fixed-effects meta-analysis. To exclude confounding due to survivorship bias, we performed the analysis using competing risks regression, with death as the competing risk. The R packages survival (https://cran.r-project.org/web/packages/survival/index.html), meta (https://cran.r-project.org/web/packages/meta/index.html), and cmprsk (https://cran.r-project.org/web/packages/cmprsk/index.html) were used to perform the Cox models, meta-analysis, and competing risks regression (CRR), respectively. Visual examination of a plot of the Schoenfeld residuals revealed that the proportional hazards assumption was met for each covariate. For FHS, some participants were selected as part of family studies, which could potentially lead to biased estimates in the regression models due to correlated genetic or environmental factors. To control for this possibility, we also included family as a cluster variable in the CRR model for FHS using the R package crrSC (https://cran.r-project.org/web/packages/crrSC/index.html) and obtained very similar results as with the un-clustered model (Table S4). Therefore, we omitted family as a variable for additional analyses.

*ADSP*

Having demonstrated a surprising inverse association between CHIP and AD dementia in the discovery set, we wished to replicate the finding. For this, we utilized the ADSP data. As described above, carriers of *APOE* ε2 or *APOE* ε4 alleles were selected in such a way that cases and controls were poorly matched for age. Due to this selection bias, carriers of these alleles were excluded from the analysis. However, *APOE* ε3ε3 carriers were well matched for age, allowing for us to use this set as the replication cohort. We further excluded those without blood as the source for DNA and those without a known age at blood draw used for sequencing. A major difference between ADSP and TOPMed is the use of higher depth whole exome sequencing in ADSP, compared to lower depth whole genome sequencing in TOPMed. In order to perform a power calculation for the replication study in ADSP, we had to ensure the variant allele fraction was comparable between ADSP and TOPMed for two reasons. First, the sensitivity to detect CHIP is linked to the sequencing depth, therefore the prevalence of CHIP was higher in ADSP. Second, the associations for previously studied health outcomes related to CHIP are dependent on clone size, with small clones having less of an effect size. We empirically determined that a cutoff of VAF at 0.08 gave a nearly identical VAF distribution for CHIP clones in ADSP as compared to TOPMed.

After these exclusions, we had 2,550 persons in ADSP for the analysis, of whom 43% were AD cases and 17% were CHIP carriers at a VAF>0.08. We then used the powerMediation (https://cran.r-project.org/web/packages/powerMediation/index.html) package in R to perform a power calculation for varying effect sizes of CHIP at an alpha of 0.1. For an odds ratio of 0.6 (similar to the hazard ratio for CHIP obtained from TOPMed), the power was 1. For an odds ratio of 0.8, the power was 0.96. For an odds ratio of 0.9, the power was 0.50. Thus, we were well-powered for the replication analysis in ADSP. We used logistic regression to assess for the association between CHIP and AD in ADSP, with age at blood draw and sex as other explanatory variables in the model. For privacy concerns, those age 90 or older did not have an exact age available on dbGaP, and were considered to be age 90 for the purposes of this analysis.

We further assessed whether smaller clones were associated with AD dementia in two ways. First, we performed a logistic regression for AD where CHIP status was modeled as a three-factor variable (no CHIP, CHIP with VAF≤0.08, or CHIP with VAF>0.08). Second, we performed a logistic regression for AD dementia where VAF was included as a continuous variable.

A fixed-effects meta-analysis for risk of AD in CHS, FHS, and ADSP was performed using logistic regression models for each cohort with age at blood draw, sex, APOE genotype and CHIP carrier status as covariates.

We wished to test whether CHIP status was associated with AD-related pathologic changes in people without clinical dementia symptoms. For a subset of participants in ADSP who died and donated their brains for research, a neuritic plaque score based on the Consortium to Establish a Registry for Alzheimer's Disease (CERAD) criteria and Braak stage was assessed. For this analysis, we utilized all *APOE* genotypes and limited the analysis to those with available age at autopsy. Information on CERAD score was obtained from the National Alzheimer's Coordinating Center Alzheimer's Disease Research Centers (NACC/ADRC) and the Adult Changes in Thought (ACT) cohort. Information on Braak stage was available from NACC/ADRC, ACT, FHS, and the Genetic Differences cohort. For all cohorts, anyone with a clinical dementia diagnosis was excluded. For NACC/ADRC and ACT, we also excluded anyone with mild cognitive impairment. A total of 427 cases had CERAD neuritic plaque scores and 454 cases had Braak stages available for this analysis after these exclusions. We performed ordinal logistic regression for CERAD score (0-3) and Braak stage (grouped as 0/I/II, III/IV, V/VI) using the polr function in the MASS package in R (https://cran.r-project.org/web/packages/MASS/index.html). Explanatory variables included CHIP, age at death, sex, and *APOE* genotype. The t-values from the ordinal logistic model were used to calculate p-values for each of the covariates using a standard normal distribution.

*Mendelian randomization*

Data from Bick et al. (*7*)was used to select variants for the CHIP instrumental variable and data from Schwartzentruber et al. (*16*) provided summary statistics from a large AD GWAS/GWAX meta-analysis. The AD GWAS included ~22,000 AD cases and ~42,000 controls from Kunkle et al.(*31*), while the GWAX contained ~53,000 AD-by-proxy cases and ~378,000 controls from UK Biobank. We defined variants as being independent if they had a linkage disequilibrium (LD) $R^2$ value of less than 0.2 in TOPMed (http://topld.genetics.unc.edu/topld/). We selected the lead variant at each independent locus, or a variant in high LD ($R^2$>0.9) with the lead variant if the lead variant was not represented in the array data from all cohorts used in the AD GWAS/GWAX. We also excluded variants from the CHIP GWAS that were primarily restricted to African ancestry, since the AD GWAS summary statistics were obtained from Europeans. We identified three loci that met these criteria, two at the *TERT* locus (rs2853677 and rs7726159) and one at the *KPNA4*/*TRIM59* locus (rs58322641). The beta values and standard errors for these three variants from the CHIP and AD GWAS summary statistics were then used to perform inverse-variance weighted Mendelian randomization using the MendelianRandomization package (https://cran.r-project.org/web/packages/MendelianRandomization/index.html).

**Nuclei isolation from human postmortem brain tissue**

ACT is a longitudinal, community-based observational study of brain aging in participants older than 65 randomly sampled from the Group Health Cooperative (now Kaiser Permanente), a health management organization in King County, Washington. A subset of participants in the study donate their brains for research upon death, and a comprehensive neuropathological exam is performed to assess for AD and related neurodegenerative disease pathologies (*32*). For decedents with post-mortem interval of less than 8 hours, a rapid autopsy is performed in which numerous samples from multiple brain regions are taken from one hemisphere and flash frozen in liquid nitrogen.

For this analysis, we obtained occipital cortex samples from 9 ACT brain donors (8 CHIP carriers and 1 non-carrier). Three of these also had a frozen sample from cerebellum available, and 2 had a frozen sample from the putamen. For nuclei isolation, we performed and adapted the protocol from (*33*). Briefly, around 250 mg of frozen postmortem brain tissue was thawed in 5 mL lysis buffer and transferred to a douncer placed on ice. After 20-30 strokes, the homogenized tissue was transferred to a clear 50 mL ultracentrifuge tube and the volume was adjusted to 12 mL. 21 mL of sucrose buffer was added to the bottom of the clear ultracentrifuge tube, to create a concentration gradient with the homogenized tissue solution on top of the sucrose buffer. The tubes were placed in buckets in a SW32Ti swinging rotor (Beckton Dickinson). The samples were ultracentrifuged at 107163.6 RCF for 2.5 hours at 4°C. The supernatant was removed and 500 μL of 1X PBS was added to the pellet and incubated for 20 min on ice. The nuclei were then resuspended and transferred into a microcentrifuge tube. The nuclei were counted using trypan blue dilution and then centrifuged at 500G for 5 min.

Lysis buffer: 0.32M Sucrose, 5 mM CaCl2, 3 mM Mg(Acetate)$_2$, 0.1 mM EDTA, 10mM Tris-HCl pH8, 1 mM DTT, 0.1%Triton X-100 in H20.

Sucrose buffer: 1.8 M Sucrose, 3 mM Mg(Acetate)$_2$, 1 mM DTT, 10 mM Tris-HCl, pH8 in H20

**Immunostaining and sorting of the nuclei**

The nuclei were resuspended at a concentration of 200,000 cells in 50ul of 0.5% BSA in 1x PBS solution and stained for 45 min with Anti-NeuN Antibody Alexa Fluor 488(EMD Millipore) at a concentration of 1: 400, and Anti-C-MAF antibody PE (BD biosciences) at a concentration of 1: 50. The nuclei were then washed and strained using a 40um strainer. The sorting was done on an Aria II sorter using a 100um nozzle. The nuclei were collected in 0.5% BSA in 1x PBS solution and centrifuged at 500G for 5 min.

**DNA extraction, amplification and sequencing**

DNA was extracted from the nuclei using the Qiagen QIAmp DNA micro kit. DNA concentration was measured using the Qubit fluorometer. PCR was performed to amplify the region surrounding the mutation of interest (around 300bp) using the Phusion high fidelity master mix (New England Biolabs). The amplified DNA was purified using the Qiagen QIAquick PCR purification kit according to the manufacturer recommendations.

Libraries were generated from the pooled amplicons using the Celero DNA-seq library kit (NuGEN). Sequencing of the libraries was performed using MiSeq Nano v2 kits. Sequencing reads were aligned with BWA (http://bio-bwa.sourceforge.net), and variant calling and annotation done with Varscan (http://varscan.sourceforge.net) and Annovar (https://annovar.openbioinformatics.org/en/latest/).

DNMT3A p.L859* : 5'- CCAGTGTTGGCTGGTGAATG-3', 5'-TAAAACCCACTGTTCCCAGGAC-3'

DNMT3A p.R635P : 5'-CAGGGTGTGTGGGTCTAGGA-3', 5'-CACACACAAGCTTCCCCTTT-3'

DNMT3A p.Y735: 5'-GGGACAGCTATTCCCGATGA-3', 5'-TAGAAGCCATTAGTGAGCTGGC-3'

SF3B1 p.H662Q : 5'-TGTCCGTAACACAACAGCTAGA-3', 5'-AGCCCAAAGGTTTGAGTCC-3'

TET2 p.C1193Y : 5'-TTGCCTCTTGAATTCATTTGCT-3', 5'-TGCCTCTTCTTAAGAGATAACACA-3'

TET2 p.KL1818fs : 5'-GGCATGTTCAACAGCTCTCT-3', 5'-ATTGACCCATGAGTTGGAGCC-3'

ASXL1 p.S444* : 5'-ACAGCGAGATGGGCATTTTA-3', 5'-CACGTGCCAAGTTGTCTGG-3'

TET2 p.A1355V : 5'-GTGTCATTCCATTTTGTTTCTGGA-3', 5'-GCTGTCCTCAGCCCAACTTA-3'

DNMT3A p.T538fs : 5'-GGAGGCCAAGGTGTGCTAC-3', 5'- GGGTCATGTCTTCAGGGCTTAG-3'

GNB1 p.K57E : 5'- AGAGCAGCCCCTGAATGTAAC-3', 5'-GTTCAGTGCAGAATGCCACA-3'

**scATAC-seq**

*Sample processing*

After nuclei isolation as described above, samples were transposed, single cells were barcoded using 10X Genomics GEMs (Gel Bead in-EMulsions), and libraries were prepared for sequencing according to the commercially available 10X Chromium™ Next GEM Single Cell ATAC Library & Gel Bead Kit v1.1. Paired end sequencing was performed on an Illumina HiSeq 2500.

*Analysis pipeline*

Fastq files were trimmed, deduplicated, filtered and aligned using the 10X cellranger-atac count pipeline, yielding a file of high quality ATAC-seq fragments for all cells per sample. Reference genome hg19 was used for compatibility with the hematopoiesis reference dataset (described below(*23*)). The fragments file for each sample was then loaded into ArchR for downstream analysis (*34*).

Cell quality control and clustering was performed using the standard ArchR pipeline. Briefly, barcodes were called as cells based on fragments per barcode and enrichment of fragments in transcription start sites (TSS) genome wide. For each sample, doublets were predicted and removed based on similarity to computationally simulated doublets. The TileMatrix and GeneScoreMatrix were computed using default settings. For the GeneScoreMatrix, imputation was performed using the ArchR implementation of MAGIC to aid visualization of the sparse ATAC-seq signals in single cells. Dimensionality reduction and clustering was performed using the TileMatrix, which tiles the genome into 500 bp windows. Although Harmony batch correction is implemented and part of the standard workflow in ArchR, we did not use any batch correction to ensure that any biological differences between the samples would be preserved. After clustering, reproducible peaks were determined for each cluster individually to ensure that cell type specific peaks were retained. Reproducible peaks for each cluster were merged into a set of disjoint, fixed width (500bp) peaks which were used to create the cell by peak matrix. ATAC-seq pseudo-bulk tracks for selected groups of cells were exported from ArchR using the `getGroupBW` function. All tracks were identically normalized using ReadsInTSS, which corrects for variation in sequencing depth and also cell quality between different groups of cells. Specific regions in the genome were visualized using the Integrative Genomics Viewer (https://software.broadinstitute.org/software/igv/).

To quantify differences between samples within cluster 9 (the analysis shown in Fig. S6C), we used `ArchR::getMarkerFeatures` for every pair of samples in this study, and also every pair of samples in the Corces 2020 data. To control for the different cell numbers within cluster 9 for the different samples, the number of cells considered (per sample) was capped using `maxCells=114`, with 114 being the lowest number of cells present across any sample (ACT6 P, Table S13). We then counted the number of peaks passing thresholds FDR < 0.1, abs(log2(fold change)) > 1 for each pairwise comparison.

*Reference datasets*

To aid in interpretation of cell types from our scATAC-seq data, we incorporated two previously published datasets (*21*),(*23*). For the Corces et. al. brain dataset, original fastq files of all 10 scATAC samples (available under GEO accession no. GSE147672) were aligned to the hg19 reference genome and then processed as described above. For the Satpathy et. al. hematopoiesis dataset, we downloaded fragments files for the 7 samples most relevant to our study, focusing on dendritic cells and monocytes. Accession numbers and sample names of these are:

GSM3722015_PBMC_Rep1_fragments.tsv.gz

GSM3722076_PBMC_Rep2_fragments.tsv.gz

GSM3722075_PBMC_Rep3_fragments.tsv.gz

GSM3722077_PBMC_Rep4_fragments.tsv.gz

GSM3722039_Dendritic_all_cells_fragments.tsv.gz

GSM3722026_Dendritic_Cells_fragments-Reformat.tsv.gz

GSM3722027_Monocytes_fragments.tsv.gz

These fragments files were processed in ArchR as described above. After clustering, we identified the predominant dendritic cell cluster and the predominant monocyte cluster based on expression of marker genes and the known sorted sample types. For example, the monocyte cluster contained nearly all cells from the monocyte sample (GSM3722027) as well as cells from the PBMC samples. Normalized bigwig files for each cluster were exported and visualized as described above.

#### Figure S1

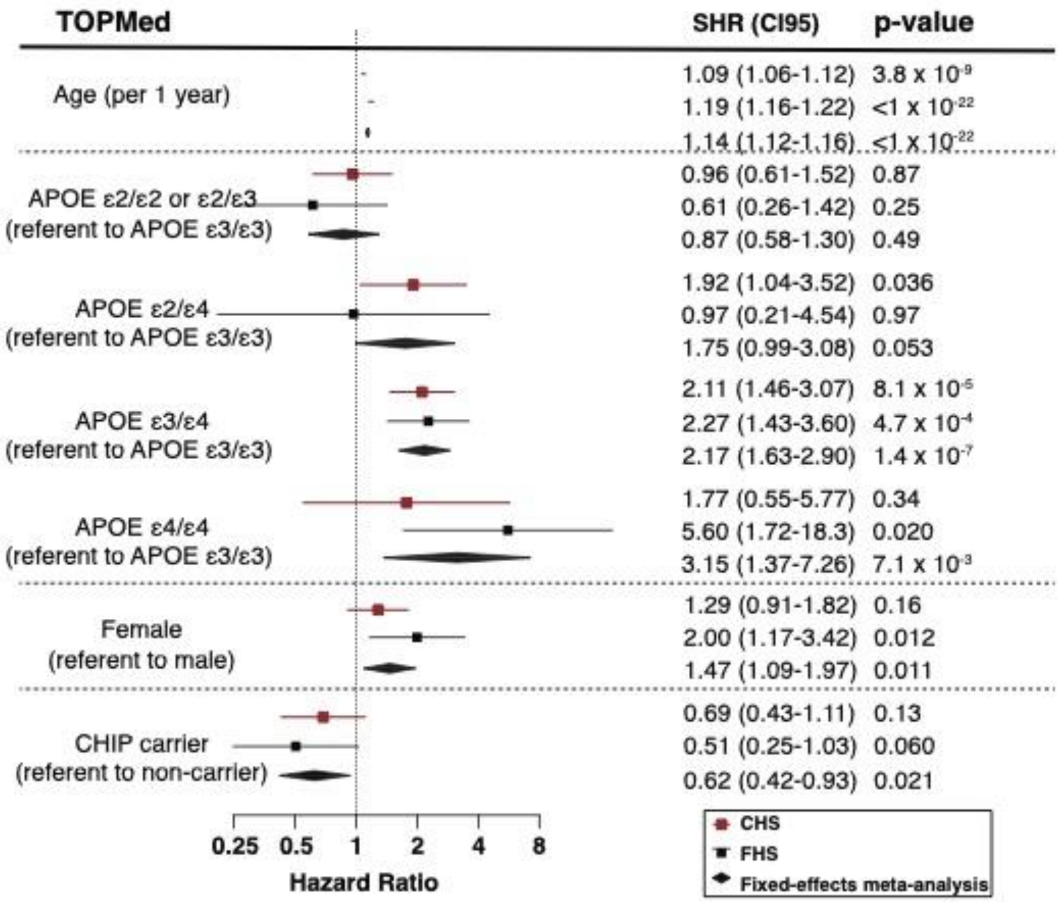


**Fig. S1. CHIP is associated with protection from AD in CHS and FHS**
Forest plot of risk factors for incident Alzheimer's disease (AD) in Cardiovascular Health Study (CHS) and Framingham Heart Study (FHS). The covariates included in the model (age at time of whole genome sequencing blood draw, *APOE* genotype, sex, and CHIP status) are shown. Hazard ratios (HR), 95% confidence intervals (CI95) and Wald p-values were calculated for each covariate from Cox proportional hazards regression models, which were then meta-analyzed using a fixed-effects model for the two cohorts.

#### Figure S2

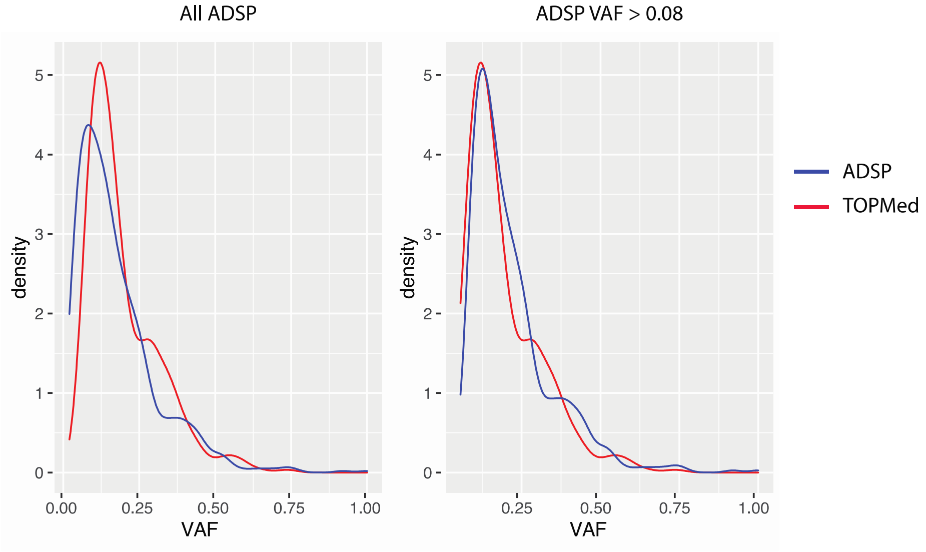


**Fig. S2. The VAF distribution is different in TOPMed and ADSP due to higher sequencing depth in ADSP** Density plots of VAF from all CHIP carriers in ADSP (left) or in CHIP carriers with VAF greater than 0.08 (right) compared to VAF distribution from TOPMed CHIP carriers (red) in each plot.

### Figure S3

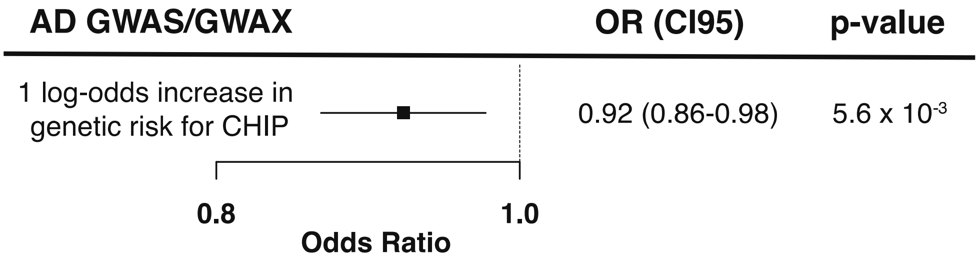


**Fig.S3. Mendelian randomization demonstrates that genetic risk of CHIP is associated with protection for AD**

Three independent genome-wide significant variants for CHIP were used to perform inverse-variance weighted Mendelian randomization on summary statistics from Schwartentruber et al. Results are presented as odds ratio (OR) for risk of AD per 1 log-odds increase in genetic risk of CHIP, with 95% confidence interval (CI95).

### Figure S4

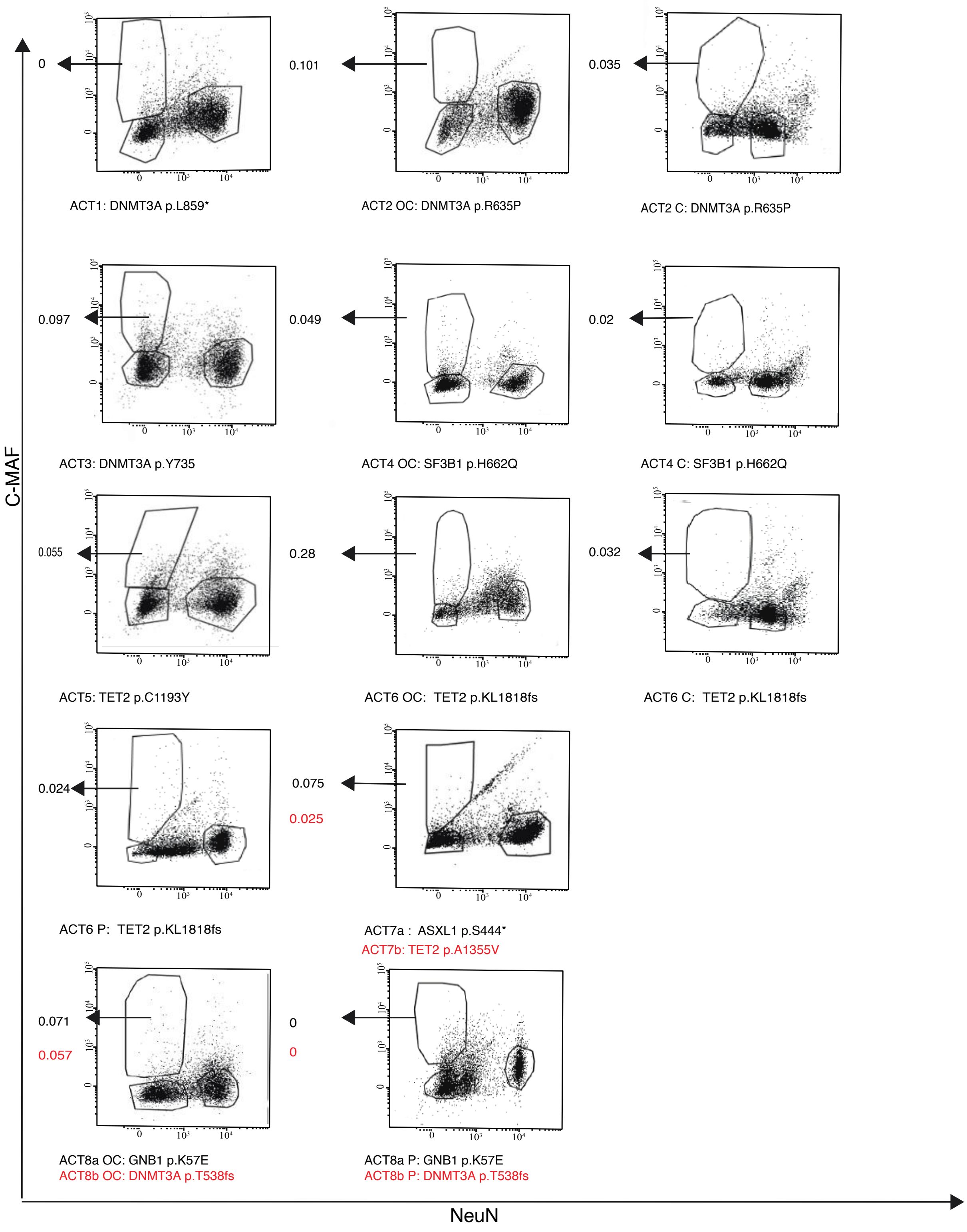


**Fig. S4. Sorting of nuclei from brains of CHIP carriers can be used to enrich for the mutant cells**
Flow cytometry gating strategy for nuclei sorting for the 8 brain samples using C-MAF and NeuN markers. The arrow points to the C-MAF$^{+}$ NeuN$^{-}$ sorted population and the VAF is indicated for each sample. The brain regions are abbreviated as Ce for Cerebellum, OC for Occipital cortex, and P for putamen.

### Figure S5

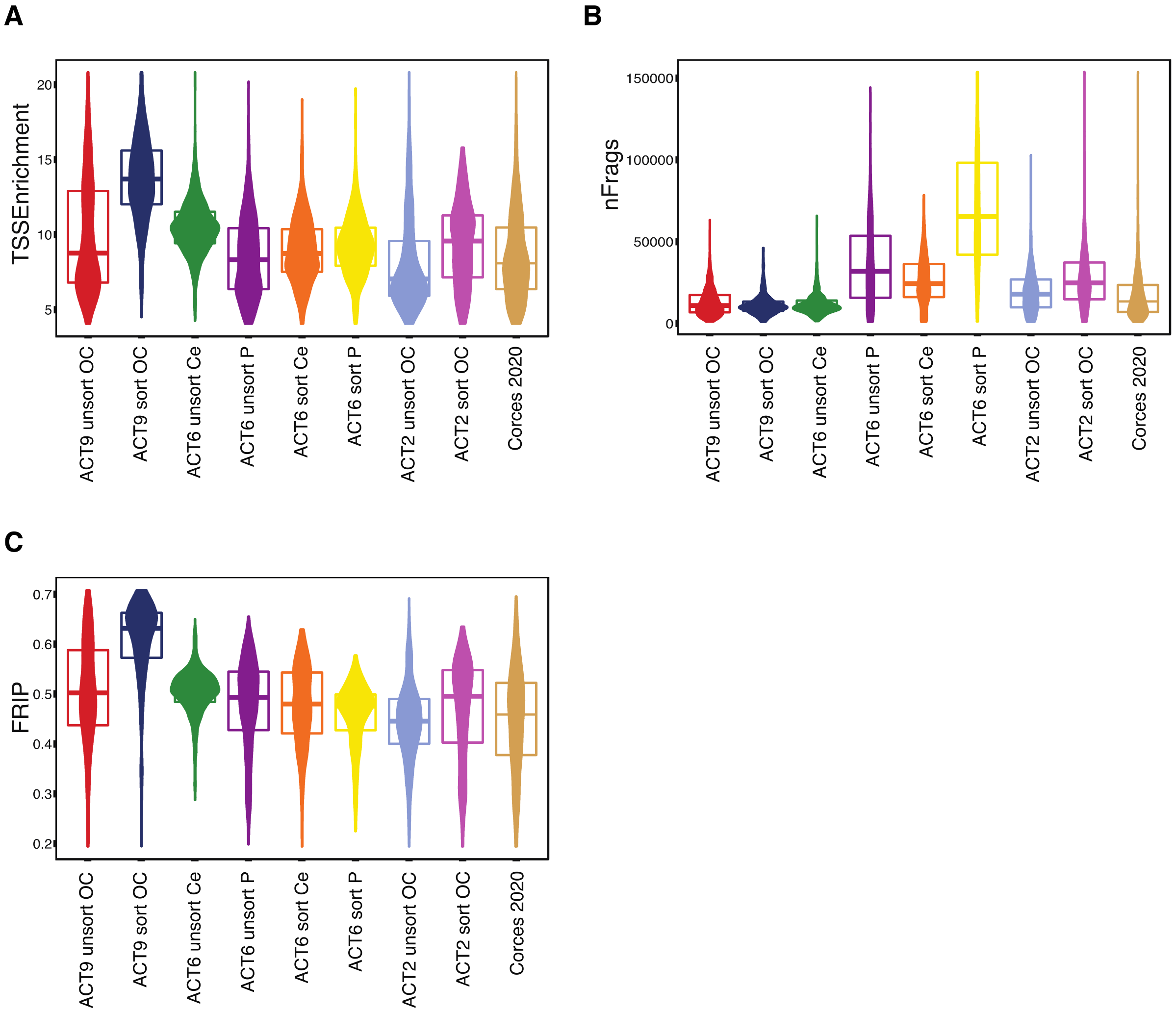


**Fig. S5. High-quality scATAC-seq libraries from ACT brain samples**
Quality control metrics for each cell in the indicated scATAC-seq sample. Aggregated Corces 2020 samples are included for reference. (A) Enrichment of fragments in transcription start sites (TSS), (B) number of fragments, and (C) fraction of reads in peaks (FRIP) for each cell.
The brain regions are abbreviated as Ce for Cerebellum, OC for Occipital cortex, and P for putamen.

### Figure S6

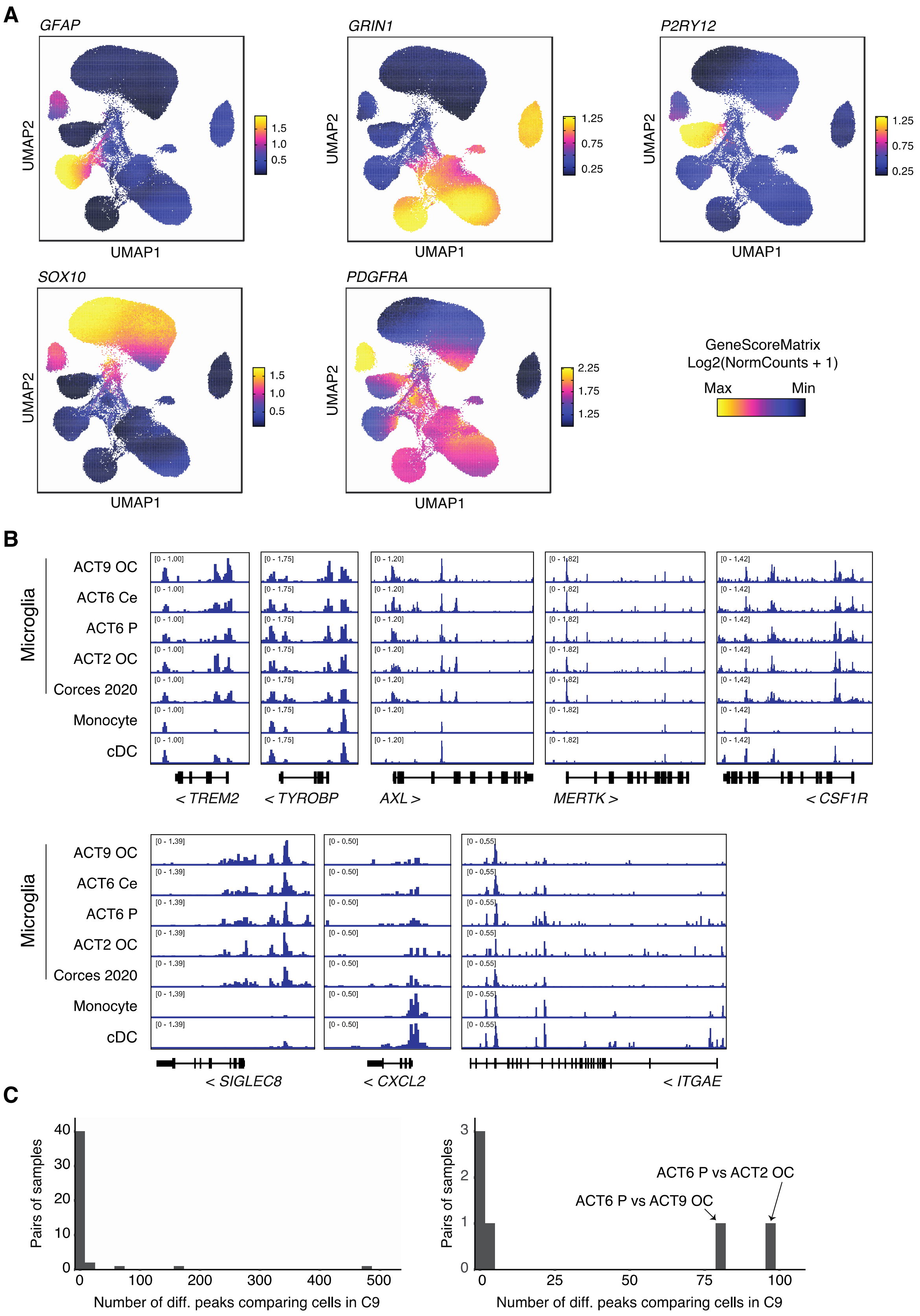
A
GFAP
GRIN1
P2RY12
SOX10
PDGFRA
UMAP1
UMAP2
GeneScoreMatrix
Log2(NormCounts + 1)
Max
Min
B
Microglia
ACT9 OC
ACT6 Ce
ACT6 P
ACT2 OC
Corces 2020
Monocyte
cDC
< TREM2
< TYROBP
AXL >
MERTK >
< CSF1R
< SIGLEC8
< CXCL2
< ITGAE
C
Pairs of samples
Number of diff. peaks comparing cells in C9
ACT6 P vs ACT2 OC
ACT6 P vs ACT9 OC

**Fig. S6. Marker genes for scATAC-seq clusters**

A) GeneScore markers used to identify cell types from scATAC-seq data.

B) Pseudo-bulk tracks shown for additional genes.

C) Histogram of the number of differential peaks (Wilcoxon test, FDR $< 0.1$, $|\log2(\text{fold change})| > 1$) comparing cells within Cluster 9 for different pairs of samples. Left: all pairs of Corces 2020 samples. Right: all pairs of samples generated in this study.

The brain regions are abbreviated as Ce for Cerebellum, OC for Occipital cortex, and P for putamen.

### Table S1. List of hematopoietic genes and variants queried

| Gene name | Reported mutations used for variant calling |
|---|---|
| ASXL1 | Frameshift/nonsense/splice-site p.358-1541 |
| ASXL2 | Frameshift/nonsense/splice-site in p.380-1435 |
| BCOR | Frameshift/nonsense/splice-site |
| BCORL1 | Frameshift/nonsense/splice-site |
| BRAF | G464E, G464V, G466E, G466V, G469R, G469E, G469A, G469V, V471F, V472S, L485W, N581S, I582M, I592M, I592V, D594N, D594G, D594V, D594E, F595L, F595S, G596R, L597V, L597S, L597Q, L597R, A598V, T599I, V600M, V600L, V600K, V600R, V600E, V600A, V600G, V600D, K601E, K601N, R603*, W604R, W604G, S605G, S605F, S605N, G606E, G606A, G606V, H608R, H608L, G615R, S616P, S616F, L618S, L618W |
| BRCC3 | Frameshift/nonsense/splice-site |
| CBL | RING finger missense p.381-421 |
| CBLB | RING finger missense p.372-412 |
| CEBPA | Frameshift/nonsense/splice-site |
| CREBBP | Frameshift/nonsense/splice-site, D1435E, R1446L, R1446H, R1446C, Y1450C, P1476R, Y1482H, H1487Y, W1502C, Y1503D, Y1503H, Y1503F, S1680del |
| CTCF | Frameshift/nonsense, R377C, R377H, P378A, P378L |
| CUX1 | Frameshift/nonsense |
| DNMT3A | Frameshift/nonsense/splice-site, F290I, F290C, F290S, G293R, L295P, L295Q, L295V, V296G, V296L, V296M, W297C, W297L, W297R, W297G, G298W, G298R, G298E, W306C, P307S, P307R, P307L, P307T, G308D, I310F, I310L, I310S, I310T, S312F, R326G, R326H, R326L, R326C, R326S, V328A, V328D, V328G, F331V, G332R, G332E, S337A, S337L, S337P, V339A, V339M, V339G, L344Q, L344P, L344R, L347P, L347R, L347Q,S352N,Y365C, R366C, R366P, R366H, R366G, A368D, A368T, A368V, I369N, I369S, V372D, L373Q, A376P, A376T, A376V, R379H, R379L, R379C, R379S, D389N, I407T, I407N, I407S, W409R, A410D, A410T, G413V, F414L, F414I, F414S, F414V, F414C, K468R, E477Q, E477K, V483G, R484Q, R484W, C494Y, C497G, C497R, C497Y, G498E, H506R, G511E, C514Y, Q527H, Q527P, D529N, D529V, D531N, D531Y, Y533C, S535F, S535P, C537G, C537R, C540Y, G543A, G543S, G543C, G543D, G543V, L547H, L547P, L547F, L547R, C537Y, M548I, M548L, M548K, M548R, M548T, G542V,G550R, C554Y, R556K, R556S, R556G, C559R, C559Y, C562Y, V563M, P580L, W581R, W581G, W581C, W581S, C583S, C583Y, C586G, C586R, C586Y, K589N, L595P, R596W, R598Q, R604Q, R604W, P633H, P633L, I634F, I634T, R635G, R635L, R635P, R635W, R635Q, V636A, V636G, V636M, V636L, L637R, L637P, L637Q, S638F, S638P, S638Y, L639R, L639V, L639F, A644T, T645A, G646V, G646E, L647H, L648P, V649G, V649L, V649M, L650V, L650Q, L653W, L653F, I655N, I655T, Q656K, V657A, V657M, V657G, D658V, D658Y, R659C, R659G, R659H, Y660C, Y660N, Y660F, Y660H, Y660D, A662D, S663L, S663W, E664K, V665G, V665L, S669F, S669P, M674V, V675A, V675M, R676L, R676W, R676Q, I681N, I681S, I681M, M682R, Y683D, V684F, G685R, G685E, G685A, D686Y, D686G, D686H, D686V, V687L, D686A, V687F, R688C, R688G, R688H, V690G, V690F, V690D, T691I, I695N, H694Y, H694P, I695F I695T, Q696P,W698C, W698R, W698S G699R, G699S, G699D, G699V, P700L, P700S, P700R, P700Q, P700T, P700A, F701V, D702A, D702G. D702E, D702V, D702N, D702Y, L703P, L703R, L703V, V704A, V704M, V704G, I705F, I705T, I705S, I705N, G706E, G706W, G706R, G706V, G707C, G707D, G707S, G707R, G707V, C710S, C710Y, D712A, L713F, S714C, V716D, V716F, V716I, N717S, N717I, P718L, R720C, R720H, R720G, R720S, K721R, K721T, K721N, Y724C, E725K, G726V, G728D, R729Q, R729W, R729G, R729L, F731C, F731L, F731Y, F731I, F731V, F732del, F732C, F732I, F732S, F732L, F732V, E733G, E733A, E733V, F734L, F734C, F734V, Y735C, Y735N, Y735S, Y735F, Y735H, R736G, R736H, R736C, R736L, R736P, R736S, L737H, L737P, L737V, L737F, L737R, L737P, L738P, L738Q, H739P, A741G, A741V, R742L, R742G, R742P, P743H, P743R, P743L, P743S, R749C, R749L, R749H, R749G, P750R, F751L, F751C, F751I, F751V, F752del, F752C, F752L, F752I, F752V, W753G, W753C, W753L, W753R, W753S, L754P, L754R, L754H, F755S, F755I, F755L, M761I, M761V, G762C, V763G, V763I, K766E, D768E, D768H, D768V, D768Y, I769N, I769S, I769T, I769V, S770L, S770W, S770P, R771G, R771L, R771P, R771Q, F772C, F772I, F772V, L773H, L773I, L773R, L773V, E774A, E774K, E774D, E774G, E774V, S775F, S775P, P777A, P777H, P777L, P777R, P777T, P777S, V778M, I780N, I780S, I780T, D781G, V785M, A787G, A787S, H789Q, R790W, A791V, R792C, R792H, R792S, F794L, F794V, W795S, W795G, W795C, W795L, W795R, G796A, G796C, G796D, G796V, N797D, N797Y, N797H, L798P, L798H, N797K, N797S, P799L, P799A, P799T, P799S, P799R, P799H, G800S, M801I, M801R, M801T, M801V, R803S, R803C, R803K, R803T, R803W, R803M, P804L, P804S, L815Q, H821D, H821P, H821R, K826N, K826T, K826R, S828N, K829R |
| EED | Frameshift/nonsense, L240Q, I363M |
| EP300 | Frameshift/nonsense, VF1148_1149del, D1399N, D1399Y, P1452L, Y1467N, Y1467H, Y1467C, R1627W, A1629V |
| ETNK1 | N244S, N244T, N244K |
| ETV6 | Frameshift/nonsense |
| EZH2 | Frameshift/nonsense, Q62R, N102S, F145S, F145C, F145Y, F145L, G159R, E164D, R202Q, K238E, E244K, R283Q, H292R, P488S, R497Q, R561H, T568I, K629E, Y641N, Y641H, Y641S, Y641C, Y641F, D659Y, D659G, V674M, A677G, A677V, R679C, R679H, R685C, R685H, A687V, N688I, N688K, H689Y, S690P, I708V, I708T, I708M, E720K, E740K |
| FLT3 | V579A, V592I, F594L, FY590-591GD, D835Y, D835H, D835E, del835 |
| GATA1 | Frameshift/nonsense |
| GATA2 | Frameshift/nonsense, R293Q, N317H, A318T, A318V, A318G, G320D, L321P, L321F, L321V, Q328P, R330Q, R361L, L359V, A372T, R384G, R384K |
| GATA3 | Frameshift/nonsense/splice-site ZNF domain, R276W, R276Q, N286T, L348V |
| GNA13 | I34T, G57S, S62F, M68K, Q134R, Y145F, L152F, E167D, Q169H, R264H, E273K, V322G, V362G, L371F |
| GNAS | R201(844)G, R201(844)S, R201(844)C, R201(844)H, R201(844)L, Q227(870)K, Q227(870)R, Q227(870)L, Q227(870)H, R374(1017)C |
| GNB1 | K57N, K57M, K57E, K57T, I80T, I80N |
| IDH1 | R132C, R132G, R132H, R132L, R132P, R132V |
| IDH2 | R140W, R140Q, R140L, R140G, R172W, R172G, R172K, R172T, R172M, R172N, R172S |
| IKZF1 | Frameshift/nonsense |
| IKZF2 | Frameshift/nonsense |
| IKZF3 | Frameshift/nonsense |
| JAK1 | T478A, T478S, V623A, A634D, L653F, R724H, R724Q, R724P, T782M, L783F |
| JAK2 | N533D, N533Y, N533S, H538R, K539E, K539L, I540T, I540V, V617F, R683S, R683G, del/ins537-539L, del/ins538-539L, del/ins540-543MK, del/ins540-544MK, del/ins541-543K, del542-543, del543-544, ins11546-547 |
| JAK3 | M511T, M511I, A572V, A572T, A573V, R657Q, V715I, V715A |
| KDM6A | Frameshift/nonsense/splice-site, del419 |
| KIT | ins503, V559A, V559D, V559G, V559I, V560D, V560A, V560G, V560E, del560, E561K, del579, P627L, P627T, R634W, K642E, V654A, V654E, H697D, E761D, K807R, D816H, D816Y, D816F, D816I, D816V, D816H, del551-559 |
| KRAS | G12D, G12A, G12E, G12V, G13D, G13C, G13Y, G13F, G13R, G13A, G13V, G13E, V14I, L19F, T58I, G60D, G60A, G60V, Q61K, Q61E, Q61P, Q61R, Q61L, Q61H, K117E, K117N, A146T, A146P, A146V |
| LUC7L2 | Frameshift/nonsense/splice-site |
| MLL | Frameshift/nonsense |
| MLL2 | Frameshift/nonsense |
| MPL | S505G, S505N, S505C, L510P, del513, W515A, W515R, W515K, W515S, W515L, A519T, A519V, Y591D, W515-518KT |
| NF1 | Frameshift/nonsense |
| NPM1 | Frameshift p.W288fs (insertion at c.859_860, 860_861, 862_863, 863_864) |

| | |
|---|---|
| **NRAS** | G12S, G12R, G12C, G12N, G12P, G12Y, G12D, G12A, G12V, G12E, G13S, G13R, G13C, G13N, G13P, G13Y, G13D, G13A, G13V, G13E, G60E, G60R, Q61R, Q61L, Q61K, Q61P, Q61H, Q61Q |
| **NXF1** | Frameshift/nonsense/splice-site |
| **PDS5B** | Frameshift/nonsense/splice-site, R1292Q |
| **PHF6** | Frameshift/nonsense/splice-site, A40D, M125I, S246Y, F263L, R274Q, C297Y, H302Y, H329L |
| **PHIP** | Frameshift/nonsense/splice-site |
| **PPM1D** | Frameshift/nonsense, exon 5 or 6 |
| **PRPF8** | M1307I, C1594W, D1598Y, D1598N, D1598V (ADD MORE VARS) |
| **PTEN** | Frameshift/nonsense, D24G, R47G, F56V, L57W, H61R, K66N, Y68H, C71Y, F81C, Y88C, D92G, D92V, D92E, H93Y, H93D, H93Q, N94I, P95L, I101T, C105F, C105S, D107Y, L112V, H123Y, C124R, C124S, K125E, A126D, K128N, R130G, R130Q, R130L, G132D, I135V, I135K, C136R, C136F, K144Q, A151T, D153Y, D153N, Y155H, Y155C, R159K, R159S, R161K, R161I, G165R, G165E, S170N, S170I, R173C, Y174D, Y177C, H196Y, R234W, G251C, D252Y, F271S, D326G |
| **PTPN11** | G60V, G60R, G60A, D61Y, D61V, D61G, Y63C, E69K, E69G, E69D, E69Q, F71L, F71K, A72T, A72V, A72D, T73I, E76K, E76Q, E76M, E76A, E76G, E139G, E139D, N308D, N308T, N339S, P491L, S502P, S502A, S502L, G503V, G503G, G503A, G503E, Q506P, T507A, T507K |
| **RAD21** | Frameshift/nonsense/splice-site, R65Q, H208R, Q474R |
| **RUNX1** | Frameshift/nonsense/splice-site, S73F, H78Q, H78L, R80C, R80P, R80H, L85Q, P86L, P86H, S114L, D133Y, L134P, R135G, R135K, R135S, R139Q, R142S, A165V, R174Q, R177L, R177Q, A224T, D171G, D171V, D171N, R205W, R223C |
| **SETBP1** | D868N, D868T, S869N, G870S, I871T, D880N, D880Q |
| **SETD2** | Frameshift/nonsense, V1190M |
| **SETDB1** | Frameshift/nonsense, K715E |
| **SF3B1** | G347V, R387W, R387Q, E592K, E622D, Y623C, R625L, R625C, R625G, N626D, R630G, H662Q, H662D, T663I, K666N, K666Q, K666T, K666E, K666R, K700E, V701F, A708T, G740R, G740E, G742D, A744P, A745P, K748E, R775P, D781G, E783K, R831Q, L833F, E862K, R957Q |
| **SFRS2** | Y44H, P95H, P95L, P95T, P95R, P95A, P107H, P95fs |
| **SMC1A** | K190T, R586W, M689V, R807H, R1090H, R1090C |
| **SMC3** | Frameshift/nonsense, R155I, Q367E, D392V, K571R, R661P, G662C |
| **SRCAP** | Frameshift/nonsense/splice-site |
| **STAG1** | Frameshift/nonsense/splice-site, H1085Y |
| **STAG2** | Frameshift/nonsense/splice-site |
| **SUZ12** | Frameshift/nonsense |
| **TET2** | Frameshift/nonsense/splice-site, missense mutations in catalytic domains (p.1104-1481 and 1843-2002), D1121Y, D1129Y, C1133R, C1133W, C1135Y, C1135F, C1135W, G1137D, G1137V, E1137K, E1137D, E1141K, E1144K, Y1148C, L1151R, G1152R, A1153T, A1153V, G1154S, C1156Y, V1157M, I1160F, I1160S, R1161G, R1161S, M1164I, E1165K, E1165D, R1167G, R1167K, R1167S, R1167M, L1172R, A1174T, I1175T, V1180D, M1185I, E1186A, G1187S, K1188R, G1192V, C1193Y, C1193W, P1194L, P1194R, I1195V, K1197E, W1198C, W1198R, V1201I, E1207D, L1209P, L1210P, C1211Y, L1212S, V1213M, V1213E, R1214W, R1214Q, R1216Q, H1219D, H1219R, H1219Y, C1221Y, C1221R, C1221S, C1221W, C1221F, L1229R, G1235E, R1235W, L1236V, A1241S, K1243R, K1243N, L1244P, Y1245C, Y1245N, L1248P, L1248R, L1252P, L1252V, G1256C, R1261C, R1261S, R1261H, R1261P, R1262W, C1263F, C1263Y, N1266D, N1266K, N1266H, N1266Y, N1266S, C1271W, C1271S, C1273S, C1273W, C1273R, Q1274P, G1275R, G1275V, G1282R, G1282D, R1283P, S1284F, F1287V, G1288D, G1288V, C1289F, C1289W, S1290L, W1291C, S1292R, M1293I, Y1294C, G1297E, G1297R, C1298Y, C1298S, K1299M, K1299Q, K1299N, F1300V, F1300L, F1300I, S1303G, S1303R, K1310Q, L1311Q, E1318E, L1322Q, L1322P, L1322R, L1326W, L1329P, L1329Q, L1332P, M1333K, L1340R, L1340P, Y1345D, Y1345C, Q1348K, Q1348R, I1349N, E1352K, A1355V, C1358S, C1358W, R1359C, R1359L, R1359P, R1359H, R1359G, R1359S, L1360R, G1361C, G1361S, G1361D, R1366H, R1366L, R1366C, P1367L, P1367S, P1367R, F1368L, G1370V, G1370R, V1371D, A1373P, A1376V, D1376G, F1377V, F1377I, C1378R, C1378Y, C1378F, H1380Y, H1380L, H1380R, H1380Q, R1380C, H1380D, H1382R, H1382P, R1383G, H1386D, R1387H, N1387S, T1393A, C1395Y, T1397I, L1398R, F1398C, F1398L, L1398H, L1398P, H1401Y, E1401A, N1403S, Q1414R, Q1414H, Q1414K, V1416I, V1417F, P1419R, D1427V, Q1435K, V1438F, G1861E, G1861R, G1861V, A1863V, H1868Y, H1868P, H1868L, G1869W, S1870L, L1872P, I1873T, I1873S, A1876T, A1876V, R1878P, R1878H, E1879Q, E1879G, E1879D, H1881N, H1881L, T1884A, T1884I, P1889L, N1890S, P1894T, P1894R, P1894L, P1894H, R1896G, I1897N, S1898F, V1900F, E1900K, V1900D, Y1902H, Y1902C, Q1903R, A1903V, H1904R, H1904Q, K1905E, T1905A, H1912R, H1912D, H1912Y, G1913D, A1919D, F1922S, Y1923H, H1925Q, K1934N, R1966C, D1981A, S1982Y |
| **TP53** | Frameshift/nonsense/splice-site, S46F, G105C, G105R, G105D, G108S, G108C, R110L, R110C, T118A, T118R, T118I, S127F, S127Y, L130V, L130F, K132Q, K132E, K132W, K132R, K132M, K132N, F134V, F134L, F134S, C135W, C135S, C135F, C135G, C135Y, Q136K, Q136E, Q136P, Q136R, Q136L, Q136H, A138P, A138V, A138A, A138T, T140I, C141R, C141G, C141A, C141Y, C141S, C141F, C141W, V143M, V143A, V143E, L145Q, W146C, W146L, L145R, V147G, P151T, P151A, P151S, P151H, P151R, P152S, P152R, P152L, T155P, T155A, V157F, R158H, R158L, A159V, A159P, A159S, A159D, A161T, A161D, Y163N, Y163H, Y163D, Y163S, Y163C, K164E, K164M, K164N, K164P, H168Y, H168P, H168R, H168L, H168Q, M169I, M169T, M169V, E171K, E171Q, E171G, E171A, E171V, E171D, V172D, V173M, V173L, V173G, R174W, R175G, R175C, R175H, C176R, C176G, C176Y, C176F, C176S, P177R, P177L, H178D, H178P, H178Q, H179Y, H179R, H179D, H179Q, R181C, R181Y, R181H, D186G, G187S, P190L, P190T, H193N, H193P, H193L, H193R, L194F, L194R, I195F, I195N, I195T, R196P, V197L, G199V, Y205D, Y205N, Y205C, V203M, Y205H, D208V, R213Q, R213P, F212I, R213L, R213Q, H214D, H214P, H214R, S215G, S215I, S215R, V216M, V217G, Y220N, Y220H, Y220S, Y220C, E224D, I232F, I232N, I232T, I232S, Y234N, Y234H, Y234S, Y234C, Y236N, Y236H, Y236C, M237V, M237K, M237I, C238R, C238G, C238Y, C238W, N239T, N239S, S241Y, S241C, S241F, C242G, C242Y, C242S, C242F, G244S, G244C, G244D, G245S, G245R, G245C, G245D, G245A, G245V, G245S, M246V, M246K, M246R, M246I, N247I, R248W, R248G, R248Q, R249G, R249W, R249T, R249M, P250L, I251N, L252P, I254S, I255F, I255N, I255S, L257Q, L257P, E258K, E258Q, D259Y, S261T, G262D, G262V, L265P, G266R, G266E, G266V, R267W, R267Q, R267P, E271K, V272M, V272L, R273S, R273G, R273C, R273H, R273P, R273L, V274F, V274D, V274A, V274G, V274L, C275Y, C275S, C275F, A276P, C277F, C277Y, P278T, P278A, P278S, P278H, P278R, P278L, G279E, R280G, R280K, R280T, R280I, R280S, D281N, D281H, D281Y, D281G, D281E, D281V, R282G, R282W, R282Q, R282P, E285K, E285V, E286G, E286V, E286K, K320N, L330R, G334V, R337C, R337L, A347T, L348F, T377P |
| **U2AF1** | D14G, S34F, S34Y, R35L, R156H, R156Q, Q157R, Q157P |
| **U2AF2** | R18W, Q143L, M144I, L187V, Q190L |
| **WT1** | Frameshift/nonsense/splice-site |
| **YLPM1** | Frameshift/nonsense/splice-site |
| **ZBTB33** | Frameshift/nonsense/splice-site, A9P, D11V, D11G, S15C, L19P, Q25E, Q25R, R26C, R26H, C32G, C32Y, V34G, V34D, T35P, R41Q, A45P, L50I, S54G, Y56C, Q59P, Q59K, V67I, V68D, L70R, R74G, I77F, I77N, Y86C, I90N, E100G, L108V, I113K, I113T, A114T, A114V, L116H, Y434H, Y434C, A435V, I437T, G438S, G438V, T441A, Y442C, D443G, I446S, P447A, L459F, E485D, Y494C, C496R, C496F, C496S, C496Y, R501T, Y503S, L509V, H512Y, H512R, N514K, S517C, Y522C, Y522H, R525C, Y526N, Y526D, C527R, C527Y, P532S, L533F, A534V, A534T, E535Q, E535K, E535A, R537C, R537H, R537L, T538A, T538R, H540N, E541K, H543D, H544R, E547K, R549K, Y550C, Q551P, C552G, C552Y, C555Y, Y562C, Q563P, S566P, Y584C, S591A |
| **ZNF318** | Frameshift/nonsense/splice-site |
| **ZRSR2** | Frameshift/nonsense, E133G, C181F, D185G, C187Y, H191Y, I202N, F239V, F239Y, N261Y, C280R, C302R, C326R, H330R |

**Table S2. TOPMed Demographics**

**A) Cardiovascular Health Study**

| | **No AD (n=577)** | **Incident AD (n=166)** |
|---|---|---|
| **Mean Age at Blood Draw ± SD** | 72.8±4.95 | 74.3±5.01 |
| **Female** | 375 (65.0) | 116 (69.9) |
| ***APOE* ε2ε2 or ε2ε3** | 101 (17.5) | 22 (13.2) |
| ***APOE* ε3ε3** | 340 (58.9) | 84 (50.6) |
| ***APOE* ε2ε4** | 17 (2.9) | 9 (5.4) |
| ***APOE* ε3ε4** | 115 (19.9) | 47 (28.3) |
| ***APOE* ε4ε4** | 4 (0.7) | 4 (2.4) |
| **CHIP carrier** | 90 (15.5) | 21 (12.7) |

**B) Framingham Heart Study**

| | **No AD (n=2345)** | **Incident AD (n=92)** |
|---|---|---|
| **Mean Age at Blood Draw ± SD** | 59.6±13.7 | 80.8±7.54 |
| **Female** | 1312 (55.9) | 73 (79.3) |
| ***APOE* ε2ε2 or ε2ε3** | 303 (12.9) | 7 (7.6) |
| ***APOE* ε3ε3** | 1532 (65.3) | 52 (56.5) |
| ***APOE* ε2ε4** | 44 (1.9) | 4 (4.3) |
| ***APOE* ε3ε4** | 430 (18.3) | 28 (30.4) |
| ***APOE* ε4ε4** | 36 (1.5) | 3 (3.3) |
| **CHIP carrier** | 143 (6.1) | 9 (9.8) |

SD—standard deviation

Parentheses indicate percent of total of no AD or incident AD groups

#### Table S3. Cox proportional hazards model for risk of AD in TOPMed

**A) Cardiovascular Health Study CPH model**

| | HR | 95% CI | p-value |
|---|---|---|---|
| **Age (per year)** | 1.12 | 1.08-1.15 | 2.8 x 10-13 |
| ***APOE* ε2ε2 or ε2ε3 (referent to APOE ε3ε3)** | 0.85 | 0.53-1.37 | 0.51 |
| ***APOE* ε2ε4 (referent to *APOE* ε3ε3)** | 2.11 | 1.04-4.24 | 0.037 |
| ***APOE* ε3ε4 (referent to *APOE* ε3ε3)** | 2.19 | 1.51-3.19 | 3.2 x 10-5 |
| ***APOE* ε4ε4 (referent to *APOE* ε3ε3)** | 1.63 | 0.58-4.57 | 0.35 |
| **Females (referent to males)** | 1.20 | 0.85-1.69 | 0.30 |
| **Davis site (referent to Bowman Gray)** | 0.87 | 0.46-1.64 | 0.67 |
| **Hopkins site (referent to Bowman Gray)** | 1.76 | 0.98-3.17 | 0.060 |
| **Pitt site (referent to Bowman Gray)** | 2.53 | 1.52-4.22 | 3.5 x 10-4 |
| **Black (referrent to white)** | 1.54 | 1.08-2.20 | 0.018 |
| **American Indian (referent to white)** | 1.43 | 0.20-10.4 | 0.72 |
| **CHIP carriers (referent to CHIP non-carriers)** | 0.73 | 0.46-1.65 | 0.19 |

**B) Framingham Heart Study CPH model**

| | HR | 95% CI | p-value |
|---|---|---|---|
| **Age (per year)** | 1.20 | 1.17-1.23 | <2 x 10-16 |
| ***APOE* ε2ε2 or ε2ε3 (referent to *APOE* ε3ε3)** | 0.68 | 0.31-1.49 | 0.33 |
| ***APOE* ε2ε4 (referent to *APOE* ε3ε3)** | 0.88 | 0.21-3.64 | 0.86 |
| ***APOE* ε3ε4 (referent to *APOE* ε3ε3)** | 2.22 | 1.40-3.52 | 7.5 x 10-4 |
| ***APOE* ε4ε4 (referent to *APOE* ε3ε3)** | 5.39 | 1.64-17.7 | 5.6 x 10-3 |
| **Females (referent to males)** | 1.66 | 0.98-2.81 | 0.062 |
| **CHIP carriers (referent to CHIP non-carriers)** | 0.57 | 0.28-1.15 | 0.12 |

**C) Meta-analysis for CHIP**

| | HR | 95% CI | p-value |
|---|---|---|---|
| **CHIP carriers (referent to CHIP non-carriers)** | 0.69 | 0.47-1.02 | 0.060 |

CPH - Cox proportional hazards
HR—hazard ratio
95% CI—95 percent confidence interval

**Table S4. Competing risks regression model for risk of AD in FHS including family as cluster variable**

| | SHR | 95% CI | p-value |
|---|---|---|---|
| **Age (per year)** | 1.19 | 1.16-1.22 | 2.8 x 10-13 |
| ***APOE* ε2ε2 or ε2ε3 (referent to APOE ε3ε3)** | 0.61 | 0.27-1.42 | 0.25 |
| ***APOE* ε2ε4 (referent to *APOE* ε3ε3)** | 0.97 | 0.20-4.64 | 0.97 |
| ***APOE* ε3ε4 (referent to *APOE* ε3ε3)** | 2.27 | 0.44-1.45 | 3.3 x 10-4 |
| ***APOE* ε4ε4 (referent to *APOE* ε3ε3)** | 5.60 | 1.89-16.57 | 1.9 x 10-3 |
| **Females (referent to males)** | 2.00 | 1.10-3.62 | 0.022 |
| **CHIP carriers (referrent to non-CHIP carriers)** | 0.51 | 0.24-1.05 | 0.068 |

#### Table S5. ADSP Demographics

**A) ADSP sample**

| **Cohort** | **n Controls (*APOE* ε3ε3)** | **n Cases (*APOE* ε3ε3)** |
|---|---|---|
| ACT | 655 | 215 |
| ADC | 441 | 516 |
| CHAP | 23 | 1 |
| CUHS | 95 | 88 |
| LOAD | 70 | 114 |
| MAP | 65 | 29 |
| NCRD | 0 | 42 |
| ROS | 87 | 49 |
| TARC | 10 | 50 |
| **Total** | **1446** | **1104** |

**B) All *APOE* ε3ε3**

| | **Controls (n=1446)** | **AD cases (n=1104)** |
|---|---|---|
| **Mean Age ± SD** | 80.5±7.07 | 80.3±6.89 |
| **Female** | 835 (57.7) | 623 (56.4) |
| **CHIP carrier (VAF>0.08)** | 240 (16.6) | 129 (11.7) |
| **CHIP carrier (VAF≤0.08)** | 60 (4.1) | 60 (5.4) |

SD - standard deviation

VAF - variant allele fraction

**Table S6. Risk of AD in ADSP**

| Logistic regression model for risk of AD | OR | 95% CI | p-value |
| --- | --- | --- | --- |
| **Age (per year)** | 1.0 | 0.99-1.01 | 0.97 |
| **Females (referent to males)** | 0.94 | 0.80-1.11 | 0.47 |
| **CHIP (referent to CHIP non-carriers)*** | 0.66 | 0.53-0.84 | 5.5 x 10-4 |

* Those with VAF≤0.08 were considered non-CHIP)

OR - odds ratio

95% CI - 95 percent confidence interval

**Table S7. Risk of AD by VAF in ADSP**

**A) Logistic regression model for risk of AD stratified by VAF cutoff**

| | OR | 95% CI | p-value |
|---|---|---|---|
| **Age (per year)** | 1.0 | 0.99-1.01 | 0.95 |
| **Females (referent to males)** | 0.94 | 0.80-1.11 | 0.49 |
| **CHIP VAF>0.08 (referent to CHIP non-carriers)** | 0.67 | 0.53-0.85 | 9.1 x 10-4 |
| **CHIP VAF≤0.08 (referent to CHIP non-carriers)** | 1.25 | 0.86-1.81 | 0.23 |

**B) Linear regression model for risk of AD in CHIP carriers using VAF as a continuous variable**

| | OR | 95% CI | p-value |
|---|---|---|---|
| **Age (per year)** | 1.0 | 0.97-1.03 | 0.80 |
| **Females (referent to males)** | 0.81 | 0.56-1.18 | 0.28 |
| **log(VAF)** | 0.78 | 0.61-0.99 | 0.045 |

OR - odds ratio

95% CI - 95 percent confidence interval

VAF - variant allele fraction

**Table S8. Neuropathological findings in ADSP controls without dementia or cognitive impairment**

**A)**

| **CERAD neuritic plaque score** | **n CHIP non-c** | **n CHIP carriers** |
|---|---|---|
| **0** | 122 | 40 |
| **1** | 106 | 20 |
| **2** | 61 | 16 |
| **3** | 59 | 3 |
| **Total** | **348** | **79** |

**B)**

| **Braak stage** | **n CHIP non-c** | **n CHIP carriers** |
|---|---|---|
| **0, I, II** | 169 | 44 |
| **III, IV** | 135 | 36 |
| **V, VI** | 63 | 7 |
| **Total** | **367** | **87** |

**C) Ordinal logistic regression model for CERAD score**

| | **OR** | **95% CI** | **p-value** |
|---|---|---|---|
| **Age at autopsy (per year)** | 1.02 | 0.99-1.05 | 0.12 |
| ***APOE* ε2ε2 or ε2ε3 (referent to *APOE* ε3ε3)** | 0.48 | 0.28-0.79 | 4.6 x 10-3 |
| ***APOE* ε2ε4, ε3ε4, or ε4ε4 (referent to *APOE* ε3ε3)** | 1.50 | 0.93-2.40 | 0.093 |
| **Females (referent to males)** | 0.98 | 0.69-1.40 | 0.92 |
| **CHIP carriers (referent to CHIP non-carriers)** | 0.50 | 0.31-0.79 | 3.2 x 10-3 |

**D) Ordinal logistic regression model for Braak stage**

| | **OR** | **95% CI** | **p-value** |
|---|---|---|---|
| **Age at autopsy (per year)** | 1.17 | 1.13-1.21 | 6.9 x 10-18 |
| ***APOE* ε2ε2 or ε2ε3 (referent to *APOE* ε3ε3)** | 1.03 | 0.61-1.74 | 0.92 |
| ***APOE* ε2ε4, ε3ε4, or ε4ε4 (referent to *APOE* ε3ε3)** | 1.41 | 0.85-2.33 | 0.18 |
| **Females (referent to males)** | 1.03 | 0.71-1.51 | 0.86 |
| **CHIP carriers (referent to CHIP non-carriers)** | 0.56 | 0.35-0.89 | 0.015 |

OR- odds ratio

95% CI - 95 percent confidence interval

CERAD - Consortium to establish a registry for Alzheimer's disease

**Table S9. Risk of AD by *APOE* genotype in TOPMed**

**A) CRR model for risk of AD in those with *APOE* ε2ε2 or ε2ε3**

| CHS | SHR | 95% CI | p-value |
|---|---|---|---|
| Age (per year) | 0.97 | 0.90-1.03 | 0.30 |
| Females (referent to males) | 0.69 | 0.30-1.57 | 0.38 |
| CHIP (referent to CHIP non-carriers) | 1.85 | 0.61-5.55 | 0.28 |
| **FHS** | **SHR** | **95% CI** | **p-value** |
| Age (per year) | 1.18 | 1.09-1.23 | 1.5 x 10-6 |
| Females (referent to males) | 1.85 | 0.16-21.5 | 0.62 |
| CHIP (referent to CHIP non-carriers) | 0.82 | 0.08-8.82 | 0.87 |

**B) CRR model for risk of AD in those with *APOE* ε3ε3**

| CHS | SHR | 95% CI | p-value |
|---|---|---|---|
| Age (per year) | 1.11 | 1.07-1.15 | 2.7 x 10-9 |
| Females (referent to males) | 1.38 | 0.85-2.23 | 0.19 |
| CHIP (referent to CHIP non-carriers) | 0.55 | 0.27-1.10 | 0.09 |
| **FHS** | **SHR** | **95% CI** | **p-value** |
| Age (per year) | 1.23 | 1.18-1.28 | <1 x 10-22 |
| Females (referent to males) | 1.66 | 0.82-3.36 | 0.16 |
| CHIP (referent to CHIP non-carriers) | 0.50 | 0.22-1.18 | 0.11 |

**C) CRR model for risk of AD in those with *APOE* ε2ε4, ε3ε4, or ε4ε4**

| CHS | SHR | 95% CI | p-value |
|---|---|---|---|
| Age (per year) | 1.05 | 1.01-1.11 | 0.032 |
| Females (referent to males) | 1.24 | 0.65-2.36 | 0.52 |
| CHIP (referent to CHIP non-carriers) | 0.71 | 0.33-1.54 | 0.39 |
| **FHS** | **SHR** | **95% CI** | **p-value** |
| Age (per year) | 1.14 | 1.10-1.18 | 1.9 x 10-14 |
| Females (referent to males) | 2.07 | 0.84-5.07 | 0.11 |
| CHIP (referent to CHIP non-carriers) | 0.29 | 0.03-2.51 | 0.26 |

CRR—Competing risks regression
SHR—Subdistribution hazard ratio
95% CI—95 percent confidence interval

**Table S10. Risk of AD by mutated gene**

**A) Logistic regression for risk of AD in TOPMed (CHS and FHS)**

| | OR | 95% CI | p-value |
|---|---|---|---|
| Age (per year) | 1.16 | 1.13-1.18 | 7.6 x 10-41 |
| Females (referent to males) | 1.70 | 1.24-2.34 | 1.0 x 10-3 |
| FHS (referent to CHS) | 0.23 | 0.17-0.31 | 2.5 x 10-22 |
| *APOE* ε2ε2 or ε2ε3 (referent to *APOE* ε3ε3) | 0.86 | 0.55-1.36 | 0.52 |
| *APOE* ε2ε4 (referent to *APOE* ε3ε3) | 1.62 | 0.76-3.46 | 0.21 |
| *APOE* ε3ε4 (referent to *APOE* ε3ε3) | 2.17 | 1.54-3.04 | 7.7 x 10-6 |
| *APOE* ε4ε4 (referent to *APOE* ε3ε3) | 4.60 | 1.79-11.8 | 1.5 x 10-3 |
| *DNMT3A* mutation (referent to CHIP non-carriers) | 0.66 | 0.34-1.29 | 0.22 |
| *TET2* mutation (referent to CHIP non-carriers) | 0.56 | 0.21-1.45 | 0.24 |
| *ASXL1* mutation (referent to CHIP non-carriers) | 0.81 | 0.21-3.13 | 0.76 |
| *SF3B1* mutation (referent to CHIP non-carriers) | 0.70 | 0.08-6.09 | 0.74 |
| Other mutation (referent to CHIP non-carriers) | 0.54 | 0.20-1.50 | 0.24 |
| Multiple mutations (referent to CHIP non-carriers) | 0.51 | 1.43-1.84 | 0.30 |

**B) Logistic regression for risk of AD in ADSP (*APOE* ε3ε3)**

| | OR | 95% CI | p-value |
|---|---|---|---|
| Age (per year) | 1.00 | 0.99-1.01 | 0.96 |
| Females (referent to males) | 0.94 | 0.80-1.10 | 0.42 |
| *DNMT3A* mutation (referent to CHIP non-carriers) | 0.75 | 0.51-1.10 | 0.14 |
| *TET2* mutation (referent to CHIP non-carriers) | 0.68 | 0.42-1.10 | 0.12 |
| *ASXL1* mutation (referent to CHIP non-carriers) | 0.55 | 0.25-1.22 | 0.14 |
| *SF3B1* mutation (referent to CHIP non-carriers) | 0.61 | 0.26-1.44 | 0.26 |
| Other mutation (referent to CHIP non-carriers) | 0.56 | 0.34-0.90 | 0.017 |
| Multiple mutations (referent to CHIP non-carriers) | 0.73 | 0.39-1.37 | 0.33 |

OR- odds ratio

95% CI - 95 percent confidence interval

**Table S11. Mutations detected from Whole Exome Sequencing of brain DNA in ADSP**

| ID | AD | *APOE* | Braak | Age | Gene | Variant Classification | Protein Change | VAF | Alt | Ref |
|---|---|---|---|---|---|---|---|---|---|---|
| ADSP_Brain_exome1 | 1 | 34 | 6 | 81-85 | *ASXL1* | Frame Shift Insertion | p.S1166fs | 0.024 | 5 | 207 |
| ADSP_Brain_exome2 | 1 | 33 | 3 | 86-90 | *BCORL1* | Nonsense Mutation | p.Y1692* | 0.103 | 3 | 26 |
| ADSP_Brain_exome3 | 1 | 33 | 5 | 86-90 | *CBL* | Missense Mutation | p.C404Y | 0.04 | 6 | 143 |
| ADSP_Brain_exome4 | 1 | 33 | 6 | 86-90 | *DNMT3A* | Missense Mutation | p.R366H | 0.086 | 5 | 53 |
| ADSP_Brain_exome5 | 1 | 34 | 5 | 76-80 | *GATA3* | Nonsense Mutation | p.C375* | 0.038 | 7 | 175 |
| ADSP_Brain_exome6 | 1 | 34 | 6 | 66-70 | *NF1* | Nonsense Mutation | p.S1786* | 0.041 | 4 | 93 |
| ADSP_Brain_exome7 | 1 | 34 | 6 | 81-85 | *NXF1* | Splice Site | | 0.06 | 5 | 79 |
| ADSP_Brain_exome8 | 0 | 33 | 1 | 86-90 | *PRPF8* | Missense Mutation | p.C1594W | 0.04 | 7 | 168 |
| ADSP_Brain_exome9 | 1 | 34 | 6 | 81-85 | *SETBP1* | Missense Mutation | p.G870S | 0.032 | 5 | 150 |
| ADSP_Brain_exome10 | 1 | 33 | 5 | 81-85 | *SRSF2* | Missense Mutation | p.P95H | 0.052 | 4 | 73 |
| ADSP_Brain_exome11 | 1 | 34 | 5 | 71-75 | *TET2* | Frame Shift Deletion | p.P656fs | 0.081 | 10 | 114 |
| ADSP_Brain_exome12 | 1 | 34 | 6 | 81-85 | *TET2* | Nonsense Mutation | p.Q810* | 0.051 | 7 | 131 |
| ADSP_Brain_exome13 | 1 | 23 | 5 | 86-90 | *TET2* | Missense Mutation | p.Q1274P | 0.068 | 4 | 55 |
| ADSP_Brain_exome14 | 1 | 23 | 5 | 86-90 | *TET2* | Nonsense Mutation | p.R1465* | 0.076 | 5 | 61 |
| ADSP_Brain_exome15 | 1 | 33 | 6 | 86-90 | *TET2* | Missense Mutation | p.R1366H | 0.058 | 6 | 97 |
| ADSP_Brain_exome16 | 0 | 33 | 0 | 86-90 | *TP53* | Missense Mutation | p.G266R | 0.089 | 24 | 247 |
| ADSP_Brain_exome17 | 1 | 34 | 5 | 76-80 | *ZBTB33* | Missense Mutation | p.Q59K | 0.042 | 4 | 92 |

AD - Alzheimer's (0 - no, 1 - yes)
VAF - variant allele fraction
Alt - alternate allele read count
Ref - reference allele read count

### Table S12. Mutations detected from sequencing of brain samples from CHIP carriers

| ID | AD | Sex | Age | *APOE* genotype | Braak stage | CHIP carrier | Gene 1 | Gene 2 | Variant 1 | Variant 2 | Blood VAF 1 | Blood VAF 2 | Region | Unsorted Brain VAF 1 | Unsorted Brain VAF 2 | c-MAF+ NeuN- VAF1 | c-MAF- NeuN- VAF1 | c-MAF- NeuN+ VAF1 | c-MAF+ NeuN+ VAF1 | c-MAF+ NeuN- VAF2 | c-MAF- NeuN- VAF2 | c-MAF- NeuN+ VAF2 | c-MAF+ NeuN+ VAF2 |
|---|---|---|---|---|---|---|---|---|---|---|---|---|---|---|---|---|---|---|---|---|---|---|---|
| ACT1 | 1 | 1 | 86-90 | 33 | 5 | yes | *DNMT3A* | | p.L859* | | 0.07 | | Occipital Cortex | <0.003 | | <0.003 | <0.003 | <0.003 | <0.003 | | | | |
| ACT2 | 0 | 1 | 86-90 | 33 | 3 | yes | *DNMT3A* | | p.R635P | | 0.14 | | Occipital Cortex | **0.0036** | | **0.101** | <0.003 | <0.003 | <0.003 | | | | |
| | | | | | | | | | | | | | Cerebellum | <0.003 | | **0.035** | | <0.003 | <0.003 | | | | |
| ACT3 | 0 | 1 | 86-90 | 33 | 2 | yes | *DNMT3A* | | p.Y735C | | 0.385 | | Occipital Cortex | **0.02** | | **0.097** | <0.003 | <0.003 | **0.0412** | | | | |
| ACT4 | 0 | 0 | 81-85 | 33 | 2 | yes | *SF3B1* | | p.H662Q | | 0.18 | | Occipital Cortex | **0.0097** | | **0.049** | **0.0073** | <0.003 | **0.0061** | | | | |
| | | | | | | | | | | | | | Cerebellum | **0.0039** | | **0.02** | **0.0071** | <0.003 | <0.003 | | | | |
| ACT5 | 0 | 1 | 86-90 | 33 | | yes | *TET2* | | p.C1193Y | | 0.111 | | Occipital Cortex | **0.0069** | | **0.055** | <0.003 | <0.003 | <0.003 | | | | |
| ACT6 | 0 | 0 | 86-90 | 33 | 2 | yes | *TET2* | | p.KL1818fs | | 0.143 | | Occipital Cortex | **0.011** | | **0.281** | <0.003 | <0.003 | <0.003 | | | | |
| | | | | | | | | | | | | | Cerebellum | **0.0016** | | **0.032** | **0.02** | <0.003 | <0.003 | | | | |
| | | | | | | | | | | | | | Putamen | **0.017** | | **0.024** | **0.015** | <0.003 | **0.0043** | | | | |
| ACT7 | 0 | 0 | 86-90 | 33 | 2 | yes | *ASXL1* | *TET2* | p.S444* | p.A1355V | 0.327 | 0.146 | Occipital Cortex | **0.0076** | <0.003 | **0.075** | <0.003 | <0.003 | <0.003 | **0.025** | <0.003 | <0.003 | <0.003 |
| ACT8 | 1 | 1 | 86-90 | 33 | 5 | yes | *DNMT3A* | *GNB1* | p.T538fs | p.K57E | 0.333 | 0.231 | Occipital Cortex | <0.003 | <0.003 | **0.071** | <0.003 | <0.003 | <0.003 | **0.057** | <0.003 | <0.003 | **0.0022** |
| | | | | | | | | | | | | | Putamen | <0.003 | <0.003 | <0.003 | <0.003 | <0.003 | <0.003 | <0.003 | <0.003 | <0.003 | <0.003 |
| ACT9 | 0 | 1 | 86-90 | 33 | 1 | no | | | | | | | Occipital Cortex | | | | | | | | | | |

AD — Alzheimer's (0 - no, 1 - yes)
Sex — 0-male, 1-female
VAF — variant allele fraction

| | C1 | C2 | C3 | C4 | C5 | C6 | C7 | C8 | C9 | C10 | C11 | C12 | C13 | C14 | C15 | C16 | C17 | C18 |
|---|---|---|---|---|---|---|---|---|---|---|---|---|---|---|---|---|---|---|
| ACT9 unsort OC | 0 | 0 | 2423 | 5776 | 17 | 0 | 5 | 1 | 211 | 39 | 1864 | 3323 | 60 | 28 | 781 | 184 | 82 | 287 |
| ACT9 sort OC | 0 | 0 | 1 | 0 | 0 | 0 | 0 | 0 | 100 | 14 | 175 | 156 | 4 | 2 | 21 | 8 | 11 | 1 |
| ACT6 unsort Ce | 8545 | 850 | 12 | 0 | 0 | 0 | 0 | 0 | 76 | 0 | 58 | 26 | 39 | 22 | 225 | 72 | 13 | 49 |
| ACT6 unsort P | 2 | 3 | 233 | 5 | 6 | 59 | 559 | 17 | 97 | 20 | 15 | 6 | 344 | 61 | 255 | 215 | 56 | 286 |
| ACT6 sort Ce | 38 | 193 | 8 | 0 | 0 | 0 | 0 | 0 | 102 | 3 | 5 | 1 | 12 | 0 | 10 | 9 | 13 | 14 |
| ACT6 sort P | 0 | 0 | 2 | 0 | 0 | 1 | 1 | 0 | 17 | 1 | 0 | 0 | 34 | 6 | 51 | 33 | 9 | 82 |
| ACT2 unsort OC | 0 | 6 | 1092 | 4814 | 28 | 7 | 1 | 5 | 159 | 105 | 842 | 311 | 159 | 183 | 357 | 294 | 863 | 292 |
| ACT2 sort OC | 0 | 0 | 9 | 7 | 0 | 0 | 0 | 0 | 43 | 6 | 29 | 1 | 7 | 5 | 58 | 10 | 31 | 37 |
| PD3_CTRL_SUNI | 0 | 0 | 297 | 38 | 27 | 0 | 0 | 0 | 652 | 15 | 780 | 102 | 2625 | 1743 | 1022 | 330 | 111 | 116 |
| PD4_CTRL_SUNI | 0 | 0 | 51 | 1 | 3 | 0 | 0 | 1 | 414 | 83 | 616 | 135 | 604 | 879 | 380 | 138 | 444 | 152 |
| PD9_CTRL_MDFG | 0 | 0 | 1035 | 1168 | 104 | 0 | 3 | 2 | 227 | 18 | 316 | 101 | 765 | 1104 | 415 | 347 | 60 | 134 |
| RCLN_CAUD_06 | 1 | 0 | 341 | 3 | 0 | 1 | 2622 | 368 | 783 | 100 | 400 | 3935 | 68 | 237 | 340 | 572 | 138 | 490 |
| RCLN_CAUD_09 | 0 | 1 | 241 | 0 | 0 | 2 | 2992 | 124 | 632 | 14 | 1039 | 2440 | 110 | 140 | 323 | 349 | 101 | 233 |
| RCLN_CAUD_14 | 0 | 0 | 159 | 1 | 4 | 2 | 1444 | 17 | 276 | 11 | 240 | 117 | 2026 | 2518 | 392 | 157 | 36 | 38 |
| RCLN_HIPP_11 | 1 | 1 | 235 | 649 | 34 | 0 | 2 | 1 | 424 | 14 | 168 | 48 | 4195 | 1112 | 387 | 171 | 36 | 147 |
| RCLN_HIPP_14 | 0 | 2 | 1018 | 113 | 882 | 15 | 8 | 6 | 890 | 17 | 589 | 400 | 462 | 2329 | 1556 | 465 | 283 | 141 |
| RCLN_PARL | 0 | 1 | 1055 | 2276 | 11 | 0 | 6 | 2 | 173 | 11 | 1210 | 471 | 252 | 45 | 63 | 131 | 18 | 49 |
| RCLN_SMTG | 0 | 3 | 1075 | 1940 | 5 | 0 | 6 | 2 | 250 | 8 | 1407 | 1141 | 92 | 34 | 53 | 188 | 18 | 51 |

**Samples in red were generated for this study. Sample in blue are from Corces et al., *Nat Gen* 2020.**

**Table S14. Calculation of percentage of mutant microglia in each sample**

| Sample | Region | Variant | Fraction of microglia in sample | VAF | Fraction of microglia with mutations |
|---|---|---|---|---|---|
| ACT9 unsorted | Occipital cortex | None | 0.013991115 | | |
| ACT9 sorted | Occipital cortex | None | 0.202839757 | | |
| ACT6 unsorted | Cerebellum | TET2 p.KL1818fs | 0.007609893 | 0.00157 | 0.411841845 |
| ACT6 sorted | Cerebellum | TET2 p.KL1818fs | 0.25 | | |
| ACT6 unsorted | Putamen | TET2 p.KL1818fs | 0.043322912 | 0.0166 | 0.766338144 |
| ACT6 sorted | Putamen | TET2 p.KL1818fs | 0.071729958 | | |
| ACT2 unsorted | Occipital cortex | DNMT3A p.R635P | 0.01670519 | 0.0036 | 0.431003774 |
| ACT2 sorted | Occipital cortex | DNMT3A p.R635P | 0.176954733 | | |
